# Insular routing to orbitofrontal cortex enables breathing awareness

**DOI:** 10.1101/2025.08.28.25334357

**Authors:** Joshua Y. Assi, Stephan Bickel, Harly E. Greenberg, Ashesh D. Mehta, Thomas Similowski, José L. Herrero

## Abstract

How does the human brain detect and respond to disruptions in breathing? While animal studies have advanced our understanding of respiratory control, breathing distress in humans remains difficult to treat. It often arises not only from pulmonary lesions or brainstem dysfunction, but also from how higher brain regions interpret breathing signals shaped by emotion and experience. We recorded intracranial cortical activity in neurosurgical patients during an interoceptive task involving transient breathing challenges. Conscious detection of these disruptions was predicted by early responses in the anterior insula, which routed signals to orbitofrontal and premotor cortices for appraisal and compensation. These cortical regions preferentially encoded inspiratory effort or airflow, revealing signal-specific processing that echoes functional segregation in brainstem centers. These findings identify a dynamic insular– frontal circuit for sensing and adapting to respiratory challenges, offering insight into the neural basis of breathing awareness and its disruption in disease.

## INTRODUCTION

Breathing is a basic life function, but its regulation depends on more than just brainstem circuits. In addition to rhythmic control by central pattern generators (1,2), higher forebrain regions actively monitor and modulate respiration (3–5). In rodents, cortical and subcortical projections to the preBötzinger complex have been identified (6,7), and manipulating these pathways alters breathing patterns—causing hyper- or hypoventilation (8,9), sniffing (10), apnea (8,11,12), or cough (13). Whether these changes generate conscious breathing sensations or dyspnea remains unknown. In human research, individuals can report such sensations, but noninvasive methods (14–17) lack the precision to track cortical processing. As a result, how the forebrain interprets internal breathing signals remains unclear: some patients fail to notice life-threatening respiratory compromise (e.g., asthmatics, opioid users, or acute COVID pneumonia (18-22)), while others report pathological breathlessness despite normal lung function (e.g., “dysfunctional breathing” associated with chronic anxiety, aging, or long COVID) (23–25). These situations likely reflect disrupted cortical processing of ascending respiratory inputs (e.g., dyspnea (26)). Understanding how the forebrain constructs conscious respiratory experience is essential to treating these disorders (27).

Leveraging rare human neurosurgical recordings and respiratory physiology, we tested the hypothesis that cortical processing of acute changes in respiratory mechanics is functionally segregated across regions. Specifically, we predicted that: (i) the insular cortex would exhibit transient, stimulus-evoked activity reflecting its role in detecting respiratory constraints; (ii) the orbitofrontal and frontal motor cortices would show more sustained responses consistent with evaluative and compensatory motor regulation; (iii) these dynamics would unfold sequentially from insular detection to frontal modulation; and (iv) inter-individual variability in detection performance would correlate with neural response strength in these regions.

To test these predictions, we directly probed respiratory perception using a novel inspiratory resistance detection task during intracranial EEG (iEEG) recordings in epilepsy patients. This approach enabled millisecond-level tracking of respiratory signal flow across multiple cortical regions involved in interoception and breathing control.

## RESULTS

To examine how the cortex detects breathing constraints, we recorded intracranial EEG (iEEG) from 11 epilepsy patients implanted with electrodes across interoceptive brain regions during a respiratory resistance sensitivity task (RRST; Fig. 1A). Three patients were excluded due to task noncompliance or excessive epileptiform activity (Table S1), yielding 8 participants and 1,328 electrodes for analysis (Fig. S1). In each trial, participants breathed through a mouthpiece with a nose clip on while brief inspiratory resistances were unpredictably applied during specific inhalations (Fig. 1B). After each trial, they reported whether they detected a load, when it occurred, and how intense it felt. Load magnitudes spanned values above and below individual perceptual thresholds, while respiratory rate and tidal volume were held constant across conditions (see Fig. S2 and supplementary text for details).

**Fig. 1.**
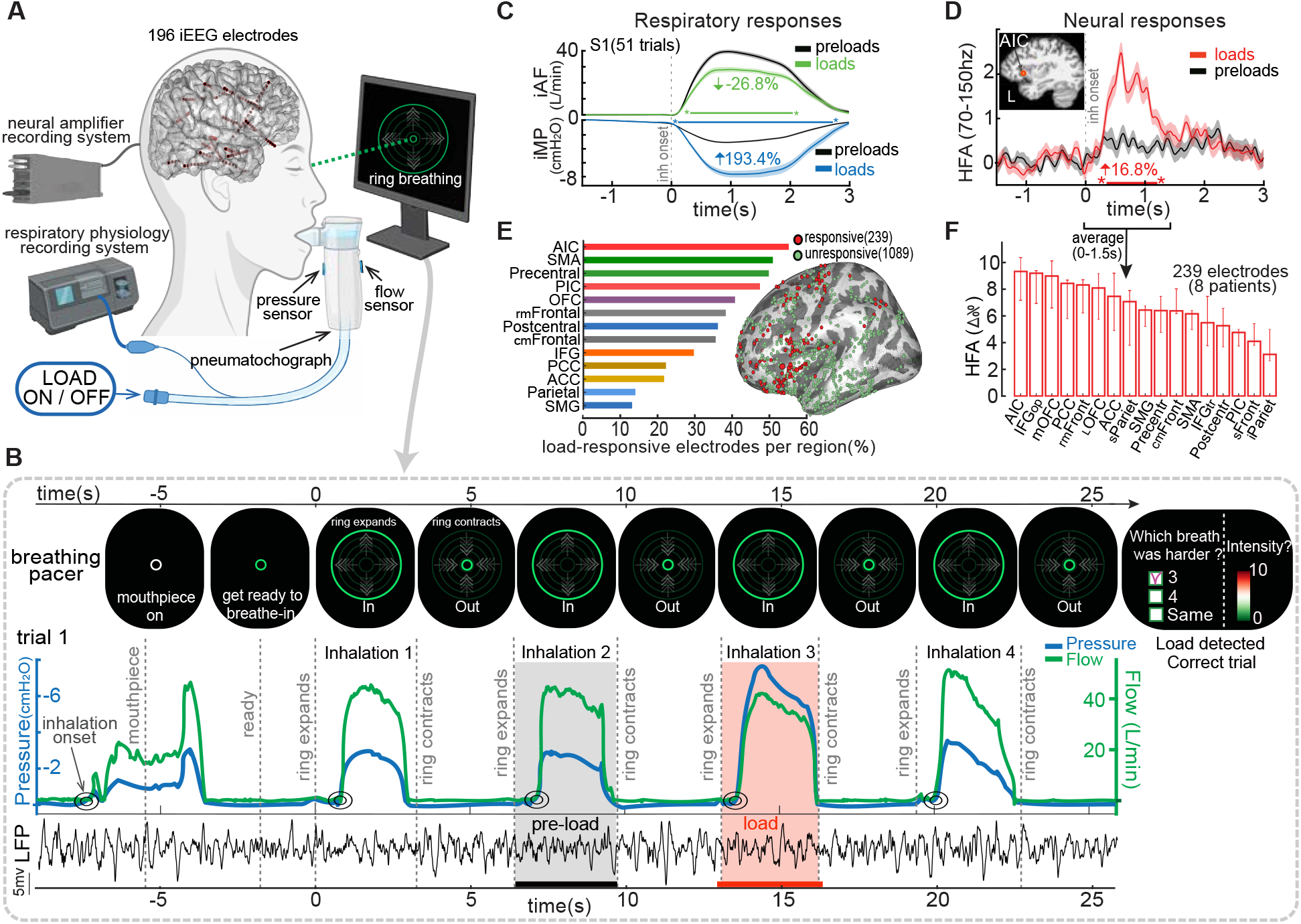
Intracranial recordings and respiratory responses during detection of inspiratory loads. **(A)** Simultaneous intracranial EEG and respiratory monitoring. Red dots show 193 sEEG electrodes implanted in cortical and subcortical regions in one patient. Patients breathed through a mouthpiece with nose clips, guided by a visual cue (expanding/contracting ring). Inspiratory loads were applied via a two-way non-rebreathing valve. Inspiratory mouth pressure (iMP) and airflow (iAF) were recorded with inline sensors. **(B)** Respiratory Resistance Sensitivity Task (RRST). Participants took four paced breaths (3 s inhale, 3 s exhale) and detected a load applied at random during one inhalation. Example trial shows effects of a 7.5 cmH2O/L/s load on iAF, iMP, and anterior insular cortex (AIC) local field potentials (LFP). **(C)** Average respiratory traces from one subject across preloaded and loaded breaths (n = 51, all load magnitudes combined). Loads decreased iAF (green) and increased iMP (red). **(D)** Example high-frequency activity (HFA; 70-150hz) from a load-responsive electrode in the AIC. **(E)** Load-responsive electrodes by brain region. Bars show the percentage of responsive electrodes relative to all recorded electrodes in each region (239/1328 total; 18%). Electrodes from the left hemisphere (all 8 participants) are overlaid on the FreeSurfer average inflated surface (see Fig. S1 for right hemisphere and alternate views). **(F)** Neural gain across regions, reflecting the strength of load-evoked HFA amplitude increases (median percent change ± IQR) during loaded vs. preloaded inhalations (0–1.5 s post-inhalation). *(sEEG, stereo-electroencephalography; LFP, local field potentials; HFA, high-frequency activity; AIC, anterior insular cortex; PIC, posterior insular cortex; mOFC, medial orbitofrontal cortex; lOFC, lateral orbitofrontal cortex; ACC, anterior cingulate cortex; PCC, posterior cingulate cortex; cmFrontal, caudomedial frontal; rmFrontal, rostromedial frontal; SMG, supramarginal gyrus; IFGop, inferior frontal gyrus pars opercularis; IFGtr, inferior frontal gyrus pars triangularis)*.

### Respiratory loads alter breathing mechanics and are reliably detected

Inspiratory loads reduced airflow (iAF) and increased mouth pressure (iMP) compared to preloaded (control) breaths (Fig. 1C). Across participants (n = 8), iAF decreased by 22.3% and iMP rose by 121.1% (p < 0.001; Fig. S3A-B), confirming effective load delivery and preserved respiratory responsiveness. Participants reliably detected suprathreshold loads, with detection declining near perceptual threshold. Mean detection rates were 8.3%, 32.2%, 48.3%, 75.9%, 87.8%, 91.1%, and 100% (chance = 33%) for load magnitudes of 1, 2.5, 5, 7.5, 10, 12.5, and 15 cmH_2_O/L/s, respectively (Fig. S3C–E). False alarms were rare (1.4%), and detection thresholds aligned with prior normative data in healthy individuals (28-29) and with normal spirometry profiles (Table S2) (30).

### Respiratory loads increase neural activity in interoceptive regions

Inspiratory loads reliably increased high-frequency activity (HFA; 70–150 Hz) relative to preloaded (control) inhalations in 18% of all electrodes (239/1,328 across 8 patients; Fig. S1). Most load-responsive sites were in insular and frontal cortices. The example AIC electrode in Fig. 1D showed a 16.8% HFA increase (0–1.5 s post-onset; p < 0.001), with effects beginning ∼285 ms after inhalation onset and lasting ∼1.5 s. Additional examples are shown in Fig. S4. The AIC had the highest proportion of responsive electrodes (53%; Fig. 1E) and the strongest HFA gains (median ΔHFA = 9.2%; p < 0.001; Fig. 1F).

### Conscious detection of respiratory loads amplifies neural responses in the anterior insula

Participants reliably detected loads ≥5 cmH_2_O/L/s (Fig. 2A), whereas lower loads were inconsistently detected near perceptual threshold (Fig. 2B, yellow box; see Fig. S3D for group data). This allowed classification of magnitude-matched load trials as detected or missed for neural comparison. In anterior insular cortex (AIC) electrodes (n = 36, 8 participants; Fig. 2C), HFA responses were notably larger during detected vs. missed trials (10.1% vs. 5.0%; p = 0.002; Fig. 2D; individual data in Fig. S5). Normalized gain ratios showed a 58% increase in AIC (Fig. 2E), with additional perception-related effects in orbitofrontal (43%) and frontal-motor regions (39%) **—**further examined in Fig. 4. Load-unresponsive electrodes (1,089/1,328) showed no such difference (p = 0.302; inset). Although inspiratory effort (iMP) was higher on detected trials, neural effects persisted after controlling for motor variability (Fig. S6).

**Fig. 2.**
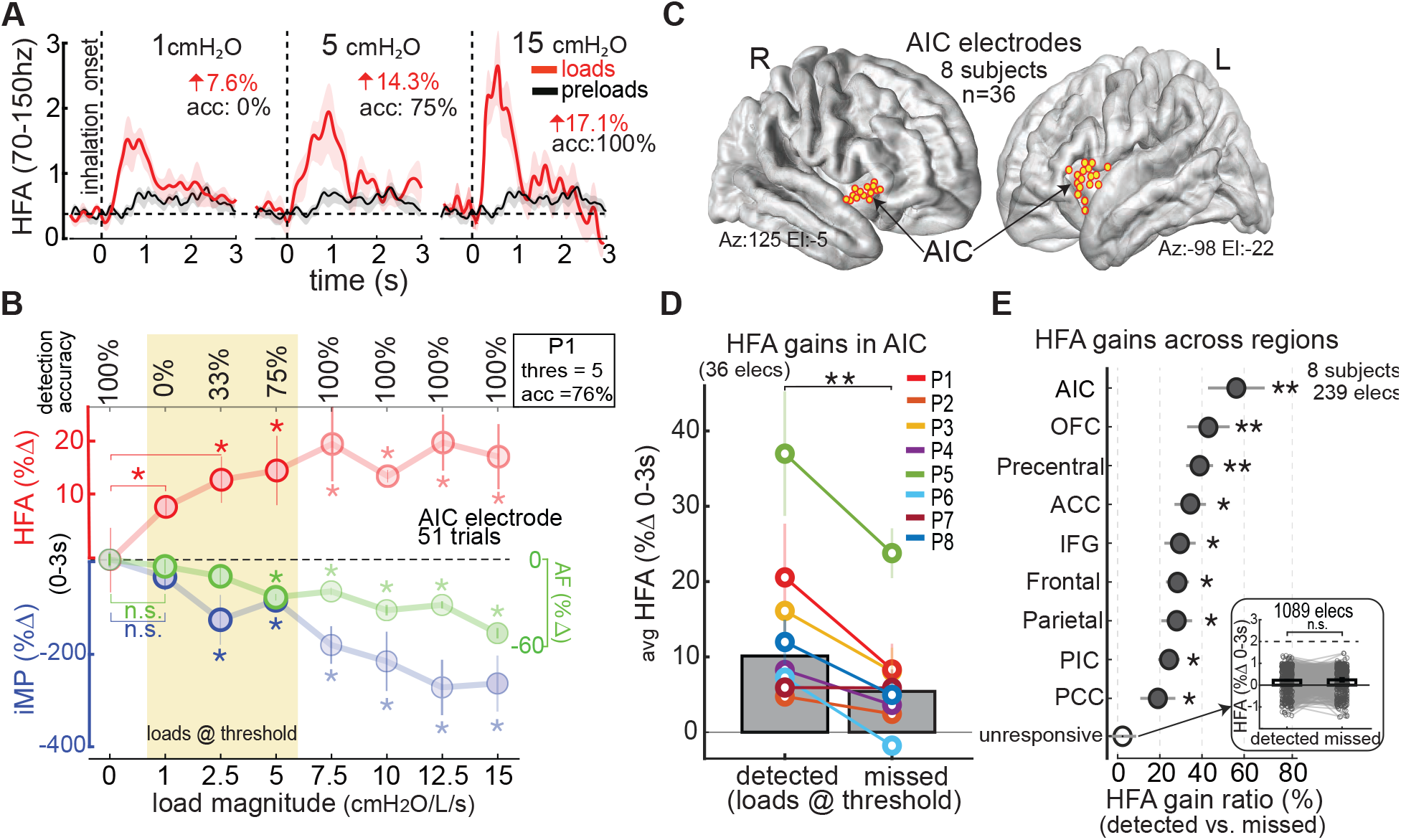
Anterior insular responses track conscious detection of inspiratory loads. **(A)** HFA responses from the same AIC electrode shown in Fig. 1D, averaged across trials for three load magnitudes (8 trials each). HFA increased with load strength. **(B)** Load-related changes in physiology and behavior from the same participant. Data show mean ± SE over a 0–3 s window. Stronger loads reduced inspiratory airflow (iAF, green) and increased mouth pressure (iMP, blue), paralleling increases in HFA (red) and detection accuracy. Yellow box marks magnitudes near perceptual threshold. Asterisks indicate significant differences from preload (load = 0); see Supplementary Text. **(C)** Load-responsive AIC electrodes (n = 36, 8 participants) plotted on the FreeSurfer average brain. **(D)** HFA gains for detected vs. missed trials across AIC electrodes, matched by load magnitude. Bars show participant means ± SE; asterisk indicates significant difference (p < 0.01, signed-rank test). **(E)** Normalized HFA gain differences across cortical regions for detected vs. missed trials. Asterisks indicate significant perception effects relative to unresponsive electrodes (^*^p < 0.05, ^**^p < 0.01). Inset shows no detection-related difference in load-unresponsive electrodes (n = 1,089). Regions with similar effects are grouped together. Precentral includes primary motor, PMC, and SMA; Parietal also includes postcentral and SMG; Frontal combines caudomedial and rostromedial areas. *(SMA, supplementary motor area; PMC, premotor cortex, SMG, supramarginal gyrus)*.

### Conscious vs. unconscious load responses

Even in missed trials with low-magnitude loads below perceptual threshold, AIC activity remained elevated, with load-evoked HFA increasing by 5% on average (Fig. 2D; see Fig. S7A). This suggests that the AIC encodes respiratory constraints even without conscious awareness. However, unlike detected trials—where HFA was modulated by the respiratory cycle—missed trials showed no such coupling, reflected by reduced phase–amplitude coupling between HFA and the phase of ongoing respiration (Fig. 3A–B). Anatomically, these unconscious responses followed a distinct distribution, with prominent gains outside the AIC, particularly in the anterior cingulate cortex (Fig. S7B). Functionally, HFA in missed trials was less sensitive to load magnitude compared to detected trials (Fig. S7C), consistent with higher-order AIC processing that integrates both stimulus intensity and perceptual awareness.

**Fig. 3.**
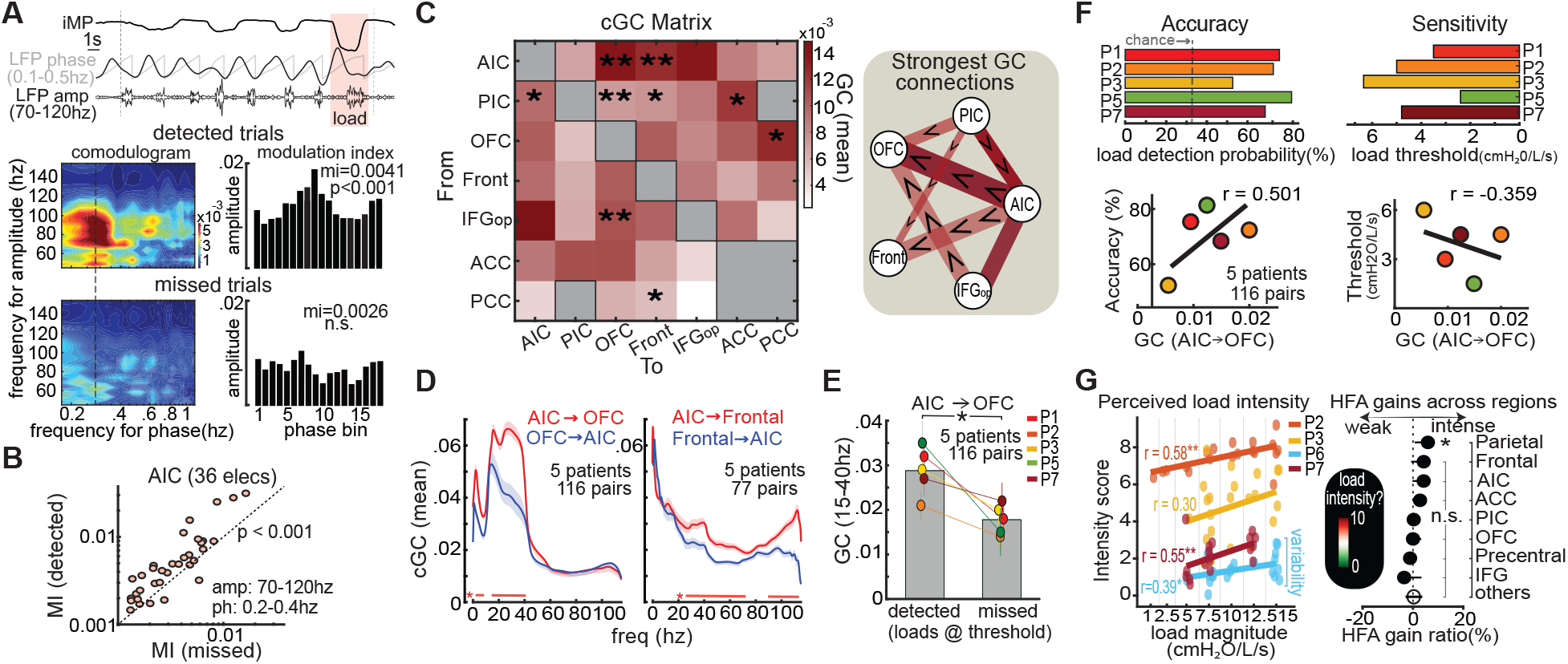
AIC coupling and network dynamics linked to conscious load detection. **A)** Phase–amplitude coupling (PAC) in an AIC electrode. Top: Example trial showing mouth pressure (iMP) and local field potential (LFP), filtered in respiratory (0.1–0.5 Hz) and high-frequency (70–150 Hz) bands. HFA amplitude tracks specific respiratory phases. Bottom: Comodulograms comparing PAC during detected and missed trials; right: phase–amplitude distributions and modulation indices (MIs). **(B)** Group PAC across AIC electrodes (n = 36, 8 participants), showing stronger HFA–respiration coupling during detected trials. **(C)** Conditional Granger causality (cGC) matrix across load-responsive regions during loaded inhalations (0–3 s post-inhalation). Darker red = stronger directed influence. Asterisks indicate significant directional asymmetries. Schematic (right) summarizes dominant connections. **(D)** Spectrally resolved GC for AIC→OFC and AIC→frontal cortex pathways (precentral, rostromedial, caudomedial). Sample sizes vary by region. **(E)** AIC→OFC GC (15–40 Hz) was stronger during detected vs. missed trials. **(F)** Left: AIC→OFC GC strength (15–40 Hz) correlated with detection accuracy. Right: Higher GC predicted lower detection thresholds. **(G) Left:** Higher inspiratory loads were rated as more intense on the visual analog scale (VAS) (n = 4 participants). **Right:** Parietal cortex HFA increased on trials rated as intense versus weak (trials were matched by their load magnitude before analyses). This effect was observed across electrodes labeled as parietal in the Desikan–Killiany atlas, including supramarginal and postcentral (sensory) regions.

**Fig. 4.**
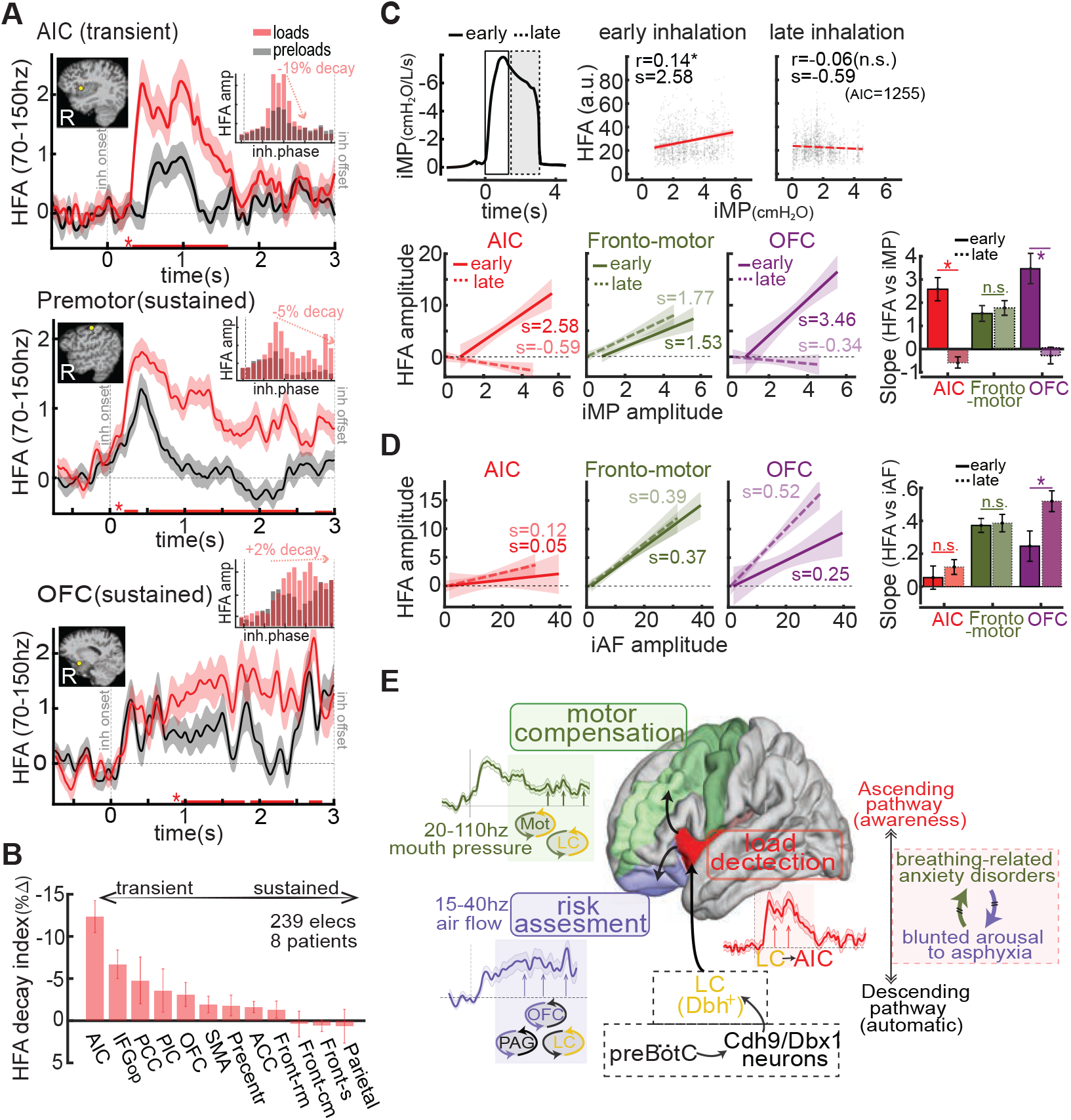
Temporal dynamics and signal-specific routing of inspiratory load to frontal cortex. **(A)** High-frequency activity (HFA, 70–150 Hz) evoked by inspiratory load in anterior insula (AIC), premotor cortex, and orbitofrontal cortex (OFC) from a representative participant (S5). Inset histograms show HFA aligned to inhalation phase, revealing a transient AIC peak and sustained frontal activity. **(B)** HFA decay index across regions (early vs. late inhalation bins). AIC showed the steepest decline, indicating transient engagement; frontal regions showed sustained or minimal decay. **(C–D)** Cortical encoding of inspiratory pressure and airflow. **(C)** Top: Example mouth pressure (iMP) trace divided into early and late phases. Right: Trial-level correlations between HFA and iMP across all AIC electrodes (dots = trials; lines = regression fits with Pearson r and slope s). Bottom: Same analysis for premotor and OFC regions. AIC responded selectively in the early phase; fronto-motor regions encoded pressure across both phases; OFC showed early-phase pressure sensitivity. **(D)** Same layout as (C), using inspiratory airflow (iAF). Fronto-motor regions showed consistent encoding; AIC responses were weak; OFC responses emerged in the late phase. Bar plots summarize group-level slope differences across regions and phases (asterisks = significant signed-rank test, FDR-corrected). **(E)** Proposed model: preBötC Cdh9/Dbx1^+^ neurons send corollary signals to the locus coeruleus (LC), which projects to the AIC for mismatch detection (expected vs. actual airflow). AIC signals reach OFC for risk evaluation and premotor cortex for motor adjustment. OFC may send descending signals to OB, PAG, and LC, forming recurrent loops that sustain appraisal. Late-phase OFC–airflow encoding suggests these loops track actual flow and update internal models, while fronto-motor regions continuously monitor pressure for motor control. Dashed boxes: hypothetical pathways (black) and affected populations (red). *(LC, locus coeruleus; PAG, periaqueductal gray; OB, olfactory bulb; preBötC, preBötzinger complex)*.

### AIC projections to OFC enable load detection

We next assessed directional cortical interactions underlying load perception using Granger causality (GC). During loaded inhalations, the AIC exerted a strong influence on the orbitofrontal cortex (OFC) and frontal motor areas (Fig. 3C). Spectral analysis revealed distinct frequency profiles: AIC→OFC connectivity peaked below 40 Hz, while AIC→frontal motor connectivity spanned 20–115 Hz (Fig. 3D). These differences may reflect divergent processing roles, possibly interoceptive salience signaling and motor compensation, respectively. Crucially, AIC→OFC connectivity increased during detected versus missed trials (p = 0.025; Fig. 3E) and correlated with both detection accuracy and sensitivity (r = 0.501 and –0.359; Fig. 3F). Other pathways were weaker and perception-insensitive, underscoring the specificity of AIC→OFC signaling in supporting conscious load detection. Additional connectivity results, including posterior insula pathways, are shown in Fig. S8.

### Perceived load intensity modulates sensory cortical responses

Participants rated perceived load intensity after each trial using a 0–10 visual analog scale (VAS; Fig. 1B). Ratings correlated with load magnitude in each of the four participants with complete data; when pooled across all trials, the correlation was r = 0.555, p = 0.017 (Fig. 3G, left). Notably, identical loads (e.g., 15 cmH_2_O/L/s) were sometimes rated as weak and other times as intense. Neural activity in the parietal, anterior insular, and frontal cortices was modestly higher during trials rated as more intense, despite matched load magnitudes. This effect reached significance only in the parietal cortex (p = 0.045; Fig. 3G, right), suggesting a role in encoding perceived load intensity.

### Load response dynamics: AIC activity is transient, frontal responses are sustained

To test whether the AIC initiates load detection while frontal regions support sustained downstream processing, we compared load-evoked HFA dynamics across regions. Example electrodes showed transient activity in the AIC and prolonged activation in frontal motor and orbitofrontal areas (Fig. 4A). To account for variability in inhalation duration (31), we normalized neural activity to the respiratory phase cycle (0–360°) and binned HFA accordingly (see insets). AIC responses were restricted to early-phase bins, whereas premotor and OFC regions remained active throughout inhalation. Across participants, HFA declined by 12% across the AIC phase cycle, but only 3% in the OFC (Fig. 4B), consistent with a temporal shift from early detection in the AIC to sustained processing in frontal cortices. Additional analyses of phase alignment, inhalation duration, and response latencies are shown in Fig. S9–S10. Moreover, the decline in AIC-to-frontal Granger causality and coherence strength at 15–50 Hz during late inhalation (Fig. S8E) suggests a shift toward recurrent brainstem–frontal cortical loops, reflecting cortico-subcortical cooperation for mechanical load compensation.

### Dissociable cortical encoding of inspiratory pressure and airflow

To identify which cortical regions encode specific features of inspiratory load, we correlated trial-level high-frequency activity (HFA) with mouth pressure (iMP) and airflow (iAF) during early and late inhalation (Fig. 4C–D; see also Fig. S11). AIC activity tracked iMP selectively during early inhalation, consistent with rapid mismatch detection of load onset. In contrast, fronto-motor regions showed sustained iMP-related modulation across both phases, and OFC also exhibited early-phase sensitivity to pressure. Slope comparisons confirmed significant regional and phase-specific differences in pressure encoding.

Airflow correlations revealed a distinct spatiotemporal profile. Fronto-motor regions again showed strong, phase-invariant coupling, while AIC was only weakly modulated by iAF. In OFC, airflow encoding emerged later, becoming prominent during late inhalation. Together, these results dissociate cortical representations of load components: the AIC preferentially signals early-phase pressure, while OFC and premotor regions integrate both pressure and airflow over time. This reinforces a functional dissociation between insular detection and frontal compensation mechanisms and suggests signal-specific organization in forebrain that mirrors brainstem tuning (e.g., airflow encoding in preBötC (1) vs. pressure/effort tracking in PAG (8)).

### Load magnitude effects

Neural responses scaled with load magnitude across regions but differed in growth profiles (Fig. 2A, B; see also Fig. S12). In the AIC, responses increased with lower loads (≤10 cmH_2_O/L/s) but plateaued or declined at higher loads. In contrast, OFC responses showed a more linear increase (Fig. S12C–D). A saturation index confirmed this pattern: AIC and pars opercularis exhibited early saturation, while other regions maintained more consistent gains (Fig. S12E). Additional magnitude effects and individual participant data are presented in Fig. S12 and supplementary text.

## DISCUSSION

We provide direct intracranial electrophysiological (iEEG) evidence for two distinct cortical populations involved in human breathing interoception (respiroception). One, in the anterior insular cortex (AIC), drives the conscious detection of subtle respiratory anomalies. This region exhibits transient, load-evoked activity consistent with early mismatch detection between expected and actual inspiratory dynamics, and relays sensory signals to downstream cortical targets (16). AIC activity alone outperformed subjective reports in identifying near-threshold loads. Notably, information flow from AIC to orbitofrontal cortex (OFC) improved load detection (Fig. 2D) and scaled with both detection accuracy and perceptual sensitivity (Fig. 3F), pointing to a cortical mechanism underlying inter-individual variability in breathing awareness.

A second population, spanning fronto-motor and orbitofrontal cortices, showed sustained load-evoked activity with distinct tuning across respiratory phases (Fig. 4). Premotor and rostromedial regions encoded both mouth pressure and airflow throughout the entire inspiration cycle, consistent with compensatory motor control and with prior EEG findings linking these regions to breathing effort (17, 32, 33). In contrast, the OFC tracked pressure early and airflow later, suggesting a shift from initial load appraisal to downstream perceptual evaluation. These dynamics support a model in which the AIC detects early deviations in expected inspiratory effort, premotor cortex mediates motor compensation, and OFC integrates the sensory and behavioral significance of the load. This cortical dissociation between inspiratory pressure and airflow echoes the functional segregation seen in brainstem centers, where regions such as the periaqueductal grey (PAG) encode effort and load perception (8, 46), while the preBötzinger complex and nucleus tractus solitarius (NTS) track airflow dynamics and timing (1, 5).

We propose that the AIC acts as a cortical gateway for ascending respiratory signals. These inputs may travel via the olfactory bulb (34–35)—unlikely here due to nasal occlusion—a somatosensory route (36), or, most plausibly, a corollary discharge pathway from brainstem centers (37, 38). This latter route likely involves excitatory input from inspiratory-active Cdh9/Dbx1^+^ neurons in the preBötzinger complex (preBötC) (9), which may be preferentially engaged during consciously detected inspiratory loads (Fig. 4E). The resulting corollary discharge could enhance mismatch detection in the AIC by flagging discrepancies between predicted and actual inspiratory effort, aligning with previous evidence implicating the AIC in error monitoring (39–41) and interoception (15, 42).

Beyond initiating detection, routing of AIC signals to the OFC appears essential for successful perception and higher-order evaluation of respiratory stimuli (Fig. 3E-F). The sustained OFC response likely reflects integration of stimulus salience or threat, maintained through recurrent interactions with brainstem centers throughout the loaded inhalation. Previous work has implicated the OFC in encoding behavioral relevance (43), including in respiratory contexts (44), and shown its direct projections to periaqueductal gray (PAG) subregions involved in respiratory control (45, 46). Through these pathways, the OFC may initiate descending loops that sustain cortical appraisal and modulate ventilation by recruiting noradrenergic (LC) and respiratory rhythm-generating (preBötC) circuits. Disruption of this AIC–OFC–brainstem network may explain not only missed detections in the RRST task but also blunted arousal responses in life-threatening conditions such as opioid-induced respiratory suppression (OIRS), sudden infant death syndrome (SIDS), COVID-19 pneumonia, and severe asthma (18–22).

Premotor and rostromedial frontal regions also showed robust load-evoked responses and received input from the AIC, notably at higher frequencies (20–110 Hz) than the AIC–OFC pathway (15–40 Hz). This spectral dissociation suggests distinct functional roles: lower-frequency AIC–OFC interactions may support sustained perceptual appraisal, while higher-frequency AIC–motor coupling likely reflects rapid motor adjustments. Prior fMRI studies demonstrate involvement of both insular and premotor cortices in response to inspiratory mechanical loads (47), and EEG studies show early supplementary motor area (SMA) activation followed by broader frontal engagement (17, 32). Our iEEG data clarify this spatiotemporal cascade, revealing strong responses in medial precentral gyrus extending into caudomedial and rostromedial frontal cortex. Notably, unlike AIC–OFC pathways, AIC–motor projections did not correlate with detection performance—suggesting these circuits support ventilatory compensation rather than conscious perception. This dissociation may help explain motor-perceptual decoupling in anxiety-related breathing disorders, such as hyperventilation without awareness (23, 24).

Our findings define a cortical AIC–OFC–motor circuit that links respiratory perception to action, forming a core substrate for interoceptive awareness and a potential target for restoring adaptive breathing control. While this circuit was delineated using transient inspiratory loads, other respiratory challenges—especially those driven by emotional or metabolic stressors—may modulate this network (70) or recruit additional regions, such as the amygdala (12, 48). By mapping these core pathways, we provide a foundation for future studies to probe how stress, arousal, and affective states shape breathing perception and drive its disruption in disease.

## MATERIALS AND METHODS

### Participants

We recorded data from neurosurgical patients with drug-resistant epilepsy undergoing intracranial EEG (iEEG) monitoring at North Shore University Hospital (New York). The study was approved by the Feinstein Institutes IRB (protocol 07-125), and all participants gave written informed consent under the Declaration of Helsinki. No compensation was provided.

Eleven patients (5 female, age 19–54 years, mean = 34.2) completed the Respiratory Resistance Sensitivity Task (RRST) (Table S1). None had a history of smoking or cardiorespiratory disease. As part of postoperative care, all were on low-dose antiepileptics and/or analgesics; testing was timed during the wear-off period to minimize drug effects.

Pulmonary function was assessed before the RRST using a hand-held spirometer, confirming normal lung function in all participants (FEV_1_, FVC ≥ 80% predicted; FEV_1_/FVC ≥ 0.70, adjusted for height) (see Table S2). BMI ranged from 19.3–29.4 kg/m^2^. During testing, patients were seated in a reclining hospital bed with support for the back, arms, neck, and head to minimize motion artifacts.

### Breathing circuit

A silicon mouthpiece (with nose clip) ensured accurate respiratory measurements, avoiding air leaks common with facemasks (30,49). Inspiratory flow was measured using a pneumotachograph (3700 series, Hans Rudolph; linear range 0–160 L/min) coupled to a 5 cmH_2_O differential pressure transducer (DP45-18, Validyne). The setup incorporated a two-way non-rebreathing valve (2700 series, Hans Rudolph). Inspiratory airway pressure was recorded via a lateral mouthpiece port connected to a separate transducer (DP15-32, Validyne; 0-140 cmH_2_O range). The inspiratory port was manually switched to a threshold-loading device just before a target inhalation (e.g., on breath #3 or #4 in each RRST trial). Expiration was unrestricted and not measured. Participants wore noise-canceling earplugs to eliminate ambient and self-generated breathing sounds. All tubing was securely fixed to prevent displacement or contact. Respiratory signals were sampled at 100 Hz using a SmartLab system (Hans Rudolph). Peak and average inspiratory flow and pressure values were extracted from a 0–3 s window after inhalation onset.

### Task and Stimuli

Stimuli were presented at bedside using the Psychophysics Toolbox v.3 in MATLAB (MathWorks). Each trial began after the participant fitted the mouthpiece and nose clip and pressed a key to display a small fixation ring on the screen (Fig. 1B). Participants fixated the ring and breathed at their natural pace and volume for 5 s (–7:–2 s). The ring then turned green (–2:0 s), signaling them to prepare to inhale. Next, the ring gradually expanded (0–3 s) and contracted (3–6 s); participants synchronized their inhalations/exhalations to match this rhythm (paced breathing: 3 s inhalation, 3 s exhalation) for four consecutive breaths. A 500 ms pause after each exhalation mimicked natural breathing.

In catch trials, no resistance was applied. In loaded trials, an inspiratory flow resistive load was randomly introduced on the 3rd or 4th breath. Participants could not see if (or when) a load was delivered. After the fourth inhalation, they removed the mouthpiece and answered: Which breath was harder? (3 vs. 4 vs. all-equal; Fig. 1B). If they reported a harder breath, they rated its intensity on a 0–10 visual analog scale (VAS), where 0 indicated a barely noticeable or very light load and 10 indicated a very strong or intense load. Each participant completed at least 38 trials (range: 38–59), receiving load magnitudes both above and below their perceptual threshold. Load levels were 1, 2.5, 5, 7.5, 10, 12.5, and 15 cmH_2_O/L/s, each presented at least four times. Loads were calibrated beforehand for linearity. Control trials without loads measured false alarms.

Participants were instructed to breathe in sync with the ring as naturally (i.e., without increasing tidal volume) and as stably (uniform across breaths) as possible (see supplementary text). This controlled respiratory rate and tidal volume, minimizing differences across participants and conditions. Because loads were relatively mild, applied to only one inspiration per trial, and no participants reported breathlessness during initial training (zero breathlessness scores), we did not collect breathlessness ratings during the main task to save time. Similar tasks have recently been used to assess respiratory sensitivity in behavioral (28) and neuroimaging studies (15).

### iEEG data acquisition

Patients were implanted with either depth electrodes (2 or 1.3 mm platinum cylinders; 4.4 or 2.2 mm center-to-center spacing; 0.8 mm diameter) or subdural strips/grids (2 or 3 mm platinum discs; 4 or 10 mm spacing; PMT Corporation), with placement determined by clinical needs. iEEG was recorded at 1.5 kHz (Tucker-Davis Technologies) or 1 kHz (XLTEK Quantum, Natus Medical) and referenced online to an electrode beneath the skull. Transistor–transistor logic pulses sent by the stimulus software were simultaneously recorded with the iEEG to align neural and respiratory data.

### Electrode localization

Electrode localization and visualization were performed in MATLAB using the iELVis toolbox (50). Before implantation, each participant underwent a T1-weighted 1 mm isometric structural MRI scan on a Siemens 3T MRI scanner. After implantation, a CT scan and a T1-weighted MRI were acquired and co-registered to the preoperative MRI using FSL’s BET (51) and FLIRT algorithms (52). This minimized localization errors from brain shift and allowed the CT to overlay precisely on the preoperative MRI. Electrode contacts were then semi-manually identified on the co-registered CT in BioImageSuite (53). Volumetric data from the preoperative T1 was processed with FreeSurfer (‘recon-all’, v6.0.0) (54). For surface visualization, depth electrodes were snapped to the nearest point on the FreeSurfer pial surface.

### Data preprocessing

We selected 11 neurosurgical patients based on intracranial electrode coverage of interoceptive and respiratory-related cortical regions, with a total of 1,768 implanted sites. Three were later excluded due to task noncompliance or excessive epileptiform activity (Table S1), yielding 8 participants (6 with depth electrodes, 2 with both depth and grid arrays). Contacts showing abnormal epileptiform activity, involvement in seizure onset zones (per clinical evaluation), or placement outside brain tissue were excluded. We also discarded electrodes fully within white matter, defined by proximal tissue density (PTD < –0.9) (55). iEEG time series were visually inspected for signal quality, and electrodes with systematic artifacts or interictal spikes (detected by an automated spike detector (56)) were removed. This process yielded 1,328 electrodes for analysis (Fig. S1).

iEEG data were notch filtered at 60, 120, and 180 Hz, bandpass filtered from 0.01–200 Hz, downsampled to 500 Hz, and re-referenced to the average across electrodes. Data were segmented into 45 s epochs containing all task events within a trial. iEEG and respiratory signals were aligned, and inhalation onsets were detected when inspiratory airflow (iAF) exceeded L/min or inspiratory mouth pressure (iMP) dropped below –0.2 cmH_2_O, accounting for load-related delays. Each inhalation was epoched from –6 to +6 s relative to onset. Epochs with transient artifacts were rejected by visual inspection before comparing preloaded and loaded inhalations.

### HFA analysis and selection of load-responsive electrodes

High-frequency activity (HFA; 70–150 Hz) was extracted as a neural response measure due to its correlation with local multiunit firing (57, 58). HFA was computed by bandpass filtering the signal into 10 Hz bands from 70–150 Hz (4th-order Butterworth, ‘filtfilt’), applying a Hilbert transform to extract the instantaneous amplitude, normalizing by the time-series mean, averaging across bands, and squaring to obtain power.

Electrodes were classified as load-responsive (Fig. S1) if their HFA was significantly higher during loaded versus preloaded inhalations within the 0–3 s post-inhalation window (p < 0.05, sign-rank test, >2%Δ; range = 38–59 trials per participant, 7 load magnitudes). HFA during preloaded inhalations (0–3 s) served as baseline, and HFA gain was computed as:

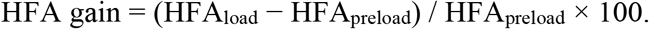

HFA gains for all electrodes within a brain region were then averaged to yield one overall HFA gain per region with p-values corrected via false discovery rate (FDR) (59).

### Perception-dependent load effects

For each participant, we identified trials where loads were applied but not detected (missed) and matched them to the nearest detected trials of the same load magnitude. HFA gains (%ΔHFA detected vs. missed, 0–3 s post-inhalation) were averaged within each electrode and brain region. These values were compared across participants using two-sided Wilcoxon signed-rank tests (Fig. 2D). HFA gain ratios were calculated for each electrode as:

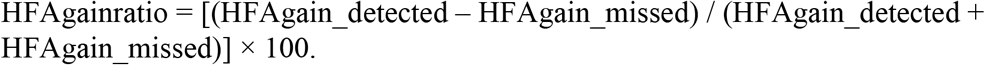

HFA gain ratios were then averaged within each region (Fig. 2E) and compared against those from load-unresponsive sites using Wilcoxon rank-sum tests, with p-values corrected for multiple comparisons via the original False Discovery Rate approach (FDR) (59). For missed trials alone (Fig. S7), HFA gains (%ΔHFA load vs. preload) were averaged across load-responsive electrodes within each region and compared to unresponsive regions. Load dependence was assessed by comparing HFA gains across load magnitudes within each region (Fig. 2B, Fig. S7C).

### Phase–amplitude coupling

We assessed how the phase of slow respiratory oscillations modulated high-frequency amplitude (HFA) in the AIC (Fig. 3A). Mouth pressure (iMP) was bandpass filtered at 0.1–0.5 Hz to extract the respiratory rhythm, and a Hilbert transform was applied to compute instantaneous phase. HFA (70–150 Hz) was extracted as described above.

We quantified phase–amplitude coupling (PAC) using the modulation index (MI) based on the Kullback–Leibler method (60). Briefly, HFA amplitudes were sorted into 18 phase bins across the respiratory cycle, and deviations from a uniform distribution were measured as MI. Comodulograms were constructed by varying phase frequencies (0.1–1.2 Hz) and amplitude frequencies (70–150 Hz) to identify peak coupling.

For group analysis (Fig. 3B), we focused on phase 0.2–0.4 Hz and amplitude 70–120 Hz, matching typical inhalation rates and broadband HFA. MIs were computed separately for detected and missed trials (matched for load magnitude) and compared within each AIC electrode using paired Wilcoxon signed-rank tests across the pooled set of electrodes (n = 36 from 8 participants).

### Granger causality

To assess the directionality of functional interactions underlying respiratory perception, we applied conditional Granger causality (cGC), which controls for shared inputs (61,62). Analyses focused on regions showing robust load responses: anterior insula (AIC), orbitofrontal cortex (OFC), pars opercularis (IFGop), frontal cortex (precentral, caudomedial, rostromedial combined), posterior insula (PIC), and anterior/posterior cingulate cortices (ACC, PCC). GC was computed on load-responsive electrode pairs within each participant, using data from loaded inhalations (0–3 s post-inhalation onset).

Local field potentials were demeaned and bipolarized by subtracting the nearest white matter signal to minimize volume conduction. To avoid errors from model order estimation, we used a non-parametric GC approach (63). Spectral pairwise-conditional GC was computed from 0–115 Hz in both forward and reverse directions. GC values were averaged within each region pair and displayed in a matrix (Fig. 3C). Significance of directional asymmetries was assessed by comparing forward vs. reverse GC with two-sided Wilcoxon signed-rank tests, correcting p-values via FDR (59).

Frequency-resolved GC profiles were averaged across electrode pairs within each connection and compared using Wilcoxon signed-rank tests (Fig. 3D). To test how GC supports conscious perception, the same analysis was repeated comparing detected vs. missed loaded trials of the same magnitude (Fig. 3E). Finally, detection accuracy and perceptual thresholds were correlated with GC values in each region using Pearson correlations (Fig. 3F).

### Transient vs. sustained responses

To capture transient versus sustained load effects, we first accounted for variability in inhalation duration *(31)* (Fig. S9A, B). We extracted inhalation phase using a Hilbert transform on an Inspiratory Efficacy (IE) signal (iAF/iMP), which provided a stable phase estimate (Fig. 4A insets, S9). Phases from −π (onset) to +π (offset) were binned into 30° segments (12 bins). Inspiratory and HFA amplitudes were averaged per bin, producing phase–amplitude distributions for loaded (red) and preloaded (black) trials (Fig. S9C, D). Bins were compared via Wilcoxon rank-sum tests, and results were aggregated across electrodes within each brain region (Fig. S9F).

To quantify suppression across the respiratory cycle, we computed an HFA Decay Index (Fig. 4B). IE signals were segmented into active, hold, and passive phases using a slope-based method (Breathmetrics toolbox, 64). Mean HFA was extracted for each phase, then fit with a linear regression. Negative slopes indicated HFA decay from early to late phases; positive slopes indicated increasing trends. These indices were averaged per electrode and then across electrodes within each region.

### Correlation analysis between HFA and inspiratory mouth pressure or airflow

To assess how cortical activity encodes mouth pressure or airflow during inspiratory load, we correlated trial-level high-frequency activity (HFA, 70–150 Hz) with inspiratory mouth pressure (iMP) or airflow (iAF) amplitudes during early (active) and late (passive) inhalation phases (Fig. 4C, D). For each electrode, we extracted per-trial HFA and iMP/iAF amplitudes and computed Pearson correlations and linear regression slopes within predefined regions of interest. Correlations and slopes were calculated separately for each phase and region. We assessed statistical significance using Pearson correlation (for within-region effects) and signed-rank tests on slope distributions (for between-region comparisons). To directly test for slope differences between regions, we fit a combined linear model with an interaction term (region × signal amplitude), treating region as a categorical predictor, using MATLAB’s fitlm function. False discovery rate (FDR) correction was applied across all tests. For visualization, regression overlays were aligned such that HFA was set to zero at the minimum iMP/iAF value, enabling direct slope comparisons. Summary panels display statistical differences in slope strength across regions and inhalation phases (see Fig. S11).

### Load magnitude responses and saturation profiles

HFA gains were computed as described above, but separately for each load magnitude (1, 2.5, 5, 7.5, 10, 12.5, and 15 cmH_2_O/L/s), yielding values such as HFAgain-5 for 5 cmH_2_O/L/s. Each participant completed at least 38 trials (mean = 42; range = 38–59), with each load presented at least four times. Gains across magnitudes were fitted with a weighted polynomial regression, assigning weights based on the standard deviation (SD) at each magnitude, to generate a load-response function (LRF) for each electrode (Fig. S12).

To quantify each electrode’s dynamic response range, we calculated a Saturation Index (SI) from the slopes between consecutive data points of the LRF, normalized by the total absolute slope (Fig. S12E). Negative slopes at higher load magnitudes were weighted more heavily to emphasize flattening. SI ranges from –1 (purely increasing response) to +1 (strong saturation), with values near 0 indicating balanced changes. Specifically,

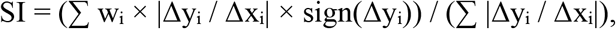

where Δy_i_ / Δx_i_ = (y_i_+1 - y_i_) / (x_i_+1 - x_i_) is the slope between consecutive points, and weights were defined as w_i_ = x_i_ / max(x). SIs were averaged across electrodes within each brain region to yield one overall SI per region.

### Load response latencies

The onset latency of HFA differences between preloaded and loaded inhalations was estimated using the phase-binned method described above (here with 24 bins for higher resolution), then mapped back to real time based on single-trial bin timing. A complementary time-domain approach, using a sliding sign-rank test (p < 0.05), produced similar results, confirming the robustness of these latency estimates. Only brain regions with consistent latency estimates across both methods and at least five electrodes were included in the summary plot (Fig. S10C).

### Statistical Analysis

Research staff were blinded to the experimental condition (load magnitude: 0, 1, 2.5, 5, 7.5, 10, 12.5, or 15 cmH_2_O/L/s) during data collection and to the detection outcome (detected vs. missed) during data preprocessing and analysis. All recorded trials from each participant were included unless excluded for technical artifacts (e.g., amplifier saturation, electrode disconnection, epileptiform discharge) or behavioral noncompliance (e.g., premature trial termination). No trials were excluded on the basis of the neural response. Participants were not randomized, as all underwent the same experimental protocol.

Sample size was determined by the availability of patients who met three criteria: (i) clinically implanted intracranial electrodes in potentially relevant interoceptive and sensorimotor regions, (ii) ability to perform the detection task (detecting at least the largest loads), and (iii) absence of excessive interictal spiking in those regions. This cohort size is consistent with prior intracranial studies of respiratory interoception (3, 48) and provides sufficient statistical power to detect large within-subject effects observed in similar designs.

When the normality assumption (Shapiro–Wilk test) was met, paired Student’s t tests were used to compare two within-subject conditions, one-way repeated-measures ANOVA to compare more than two conditions, and two-way repeated-measures ANOVA to adjust for additional within-subject factors (e.g., respiratory phase). Nonparametric counterparts (Wilcoxon signed-rank test, Friedman test) or the rank-sum test for unequal sample sizes were applied when the normality assumption was violated. Pearson correlation coefficients were used to assess linear relationships between variables; significance values were computed using two-tailed tests. Effect sizes were expressed as percent change from baseline (preload), unless otherwise noted. For cross-electrode and cross-participant comparisons, responses were normalized by computing ratios relative to the baseline condition. False discovery rate (FDR) correction was applied to account for multiple comparisons, as specified in the figure legends. For a supplementary analysis (Fig. S12), gains across load magnitudes were fitted with weighted polynomial regression to generate a load–response function for each electrode.

## Data Availability

All data produced in the present study are available upon reasonable request to the authors

## Acknowledgments

We thank Aaron Dirk from Hans Rudolph Inc. for technical support with the respiratory physiology setup as well as Tucker-Davis Technologies for assistance with neurophysiological recordings. Dr. Nima Mesgarani is gratefully acknowledged for data analyses code sharing and equipment donations.

## Author contributions

Conceptualization: JLH

Methodology: JLH, TS, HG, JYA, SB, ADM

Investigation: JLH, JYA

Visualization: JLH, JYA, TS, SB, ADM

Funding acquisition: JLH

Project administration: JLH

Supervision: JLH, JYA, TS, HG, ADM, SB

Writing – original draft: JLH, JYA

## Data and materials availability

Data and code will be made publicly available upon publication.

## Supplementary Materials

Supplementary Text

Figs. S1 to S12

Tables S1 to S2

