## Supplementary Materials for "Insular routing to orbitofrontal cortex enables breathing awareness"

**The PDF file includes:**

Materials and Methods
Supplementary Text
Figs. S1 to S12
Tables S1 to S2
References 1 to 70

**Other Supplementary Materials for this manuscript include the following:**

Movies S1 to S#
Audio S1 to S#
Data S1 to S#

### Supplementary Text

#### 1. Advantages of the Respiratory Resistance Sensitivity Task (RRST)

Previous intracranial studies examined respiration–neural correlations but did not directly probe the respiratory system (3,23,72). The RRST introduces calibrated inspiratory resistance to elicit measurable effort and directly assess respiratory interoception.

The RRST builds on prior behavioral (28) and imaging (15) paradigms by extending each trial to four paced breaths (rather than two) and adding a preparatory fixation period after mouthpiece insertion. This helps stabilize tidal volume, which typically normalizes after 1–2 breaths, and reduces load predictability—resistance was applied on the third, fourth, or no breath—enhancing perceptual sensitivity. For physiological analyses, loaded breaths were compared to the immediately preceding preload. Control analyses using matched breath positions (e.g., breath #3 across trials) produced similar results (data not shown).

#### 2. Paradigm specificity and generalization

The RRST paradigm used a metronome to maintain consistent respiratory timing and encouraged stable tidal volume through paced, even breathing. However, this setup involved voluntary breathing control and biofeedback, which may have heightened respiratory awareness or facilitated compensatory motor responses. These features—while essential for isolating load-related effects—differ from spontaneous breathing and may shape cortical engagement in ways that limit direct generalization to naturalistic conditions.

Our findings are specific to transient inspiratory loading—a brief, mechanically induced perturbation of breathing. While this approach captures both consciously detected and undetected events, the dynamics observed may not fully extend to other respiratory disruptions such as sustained mechanical loads, hypercapnia, hypoxia, or dyspnea driven by emotional or metabolic factors. These conditions may engage distinct time constants and afferent pathways (e.g., chemosensory, limbic), potentially altering network dynamics or dominant cortical and subcortical nodes. The anterior insula–frontal circuit characterized here is particularly suited to detecting and evaluating mechanical mismatch, but extrapolation to broader interoceptive or homeostatic domains should be done with caution.

#### 3. GC–Behavior Correlations

Figure 3F (left) shows that detection accuracy positively correlated with AIC→OFC cGC strength in the 15–40 Hz band (Pearson $r = 0.501$; $n = 5$ participants with AIC and OFC coverage). Bayesian estimation yielded a posterior mode of 0.622 with moderate negative skew (–0.727), reflecting some uncertainty due to small sample size. Conversely, Figure 3F (right) shows that perceptual threshold negatively correlated with AIC→OFC cGC ($r = –0.357$; posterior mode = –0.424; skew = 0.698). GC–behavior correlations in other pathways were weaker and non-significant (data not shown).

#### 4. Load response dynamics and breath-to-breath variability

Granger causality analyses showed that the AIC leads frontal regions, pointing to a role in early load detection, while frontal areas may support compensatory responses (e.g., ventilatory-motor or risk assessment). To test this, we compared load-evoked HFA dynamics across regions (Fig. 4A), accounting for breath-to-breath variability and differences in inhalation duration (31). Across participants, mean inhalation duration was 3.07 ± 0.61 s (CI: 2.75–3.40), closely matching the paced metronome (Fig. S9A–B). Individual durations ranged from 1.15–5.10 s and were slightly shorter in preload (2.97 ± 0.58 s) than load trials (3.17 ± 0.66 s; $p = 0.03$) (see Fig. S10A–B). To stabilize phase alignment, we combined airflow (iAF) and mouth pressure (iMP) into a single inspiratory efficacy index (iEff = iAF/iMP), which was then normalized from 0° to 360° (12 bins). Figure S9C shows binned iEff across all trials ($n = 339$). Example electrodes (Fig. S9E, from S9D and 4A) illustrate these dynamics: AIC activity rose in 5 of 12 bins, while premotor cortex activity remained elevated throughout inhalation. At the population level (Fig. S9F), AIC HFA decayed by -12.2% across the phase cycle, compared to weaker decay in OFC (-3%) and minimal change in other frontal regions (Fig. 4C).

Latency analyses revealed that load-evoked HFA emerged earliest in precentral regions, followed closely by the AIC (Fig. S10C), suggesting rapid preparatory engagement in both motor and insular areas. Despite this temporal proximity, Granger causality confirmed that the AIC exerted dominant directional influence on frontal circuits, supporting a model in which insular signals drive salience-based detection and evaluation even as motor regions rapidly initiate compensatory adjustments to breathing. Such dissociations—where onset latency does not dictate causal influence—have been reported in other cortical systems (66-68). For example, in visual cortex, Granger causality analyses revealed that higher-order areas (e.g., V4, TEO) exert feedback control over earlier regions despite their later response latencies (68).

**5. Load-magnitude dependent effects and response saturation**

Stronger inspiratory loads produced larger physiological and neural responses, but scaling varied by brain region. In an example AIC electrode (Fig. S12A from 2B), 1 $cmH_2O/L/s$ loads reduced iAF by just 0.75% ($p = 0.54$) yet increased HFA by 9.2% ($p < 0.001$), whereas 15 $cmH_2O/L/s$ loads reduced iAF by 52.6% and increased HFA by 17.5% (both $p < 0.001$). Notably, HFA gains plateaued or declined beyond ~10 $cmH_2O/L/s$, despite continued rises in iMP (–265%, $p < 0.001$).

To quantify these relationships, we fit load–response functions (LRFs) using curve fits similar to approaches for contrast response functions (CRFs) in primary visual cortex (69). Example AIC LRFs confirmed this saturation pattern: HFA rose steeply up to ~10 $cmH_2O/L/s$, then flattened or declined (Fig. S12B). Despite some inter-participant variability, this trend held across the AIC population (Fig. S12C). In contrast, OFC responses scaled more linearly across the full range (Fig. S12D). Saturation indices captured this divergence: AIC and IFGop showed the strongest early saturation, while other frontal, parietal, and cingulate regions exhibited steadier scaling (Fig. S12E). Across all load-responsive sites passing goodness-of-fit (231/239 electrodes), most LRFs plateaued or declined at higher loads (Fig. S12F), indicating nonlinear scaling as a general property of cortical load-responsive regions, despite differences in saturation profiles across areas (AIC > frontal).

While it would have been ideal to separately model load–response slopes for detected versus missed trials (Fig. S7C), the limited number of trials per load level precluded this. As the first iEEG study of its kind—using a design optimized to minimize expectancy and variability—each trial was necessarily lengthy, and most patients fatigued after ~40–50 trials. Addressing finer questions like detection-dependent load tuning will therefore require future paradigms tailored for higher trial throughput.

**6. Amygdala coverage and interpretation**

The amygdala was not a focus of this study due to limited electrode coverage and a lack of consistent load-evoked responses in the available sites. However, the amygdala's established role in fear-related respiratory suppression, emotional modulation of respiratory drive, and apnea warrants acknowledgment. Its omission here does not exclude its involvement in other paradigms (e.g., threat-induced breath-holding, affective dyspnea). Future work with targeted sampling in the amygdala and limbic-affiliated regions may clarify how emotional and affective load dimensions interact with mechanosensory detection.

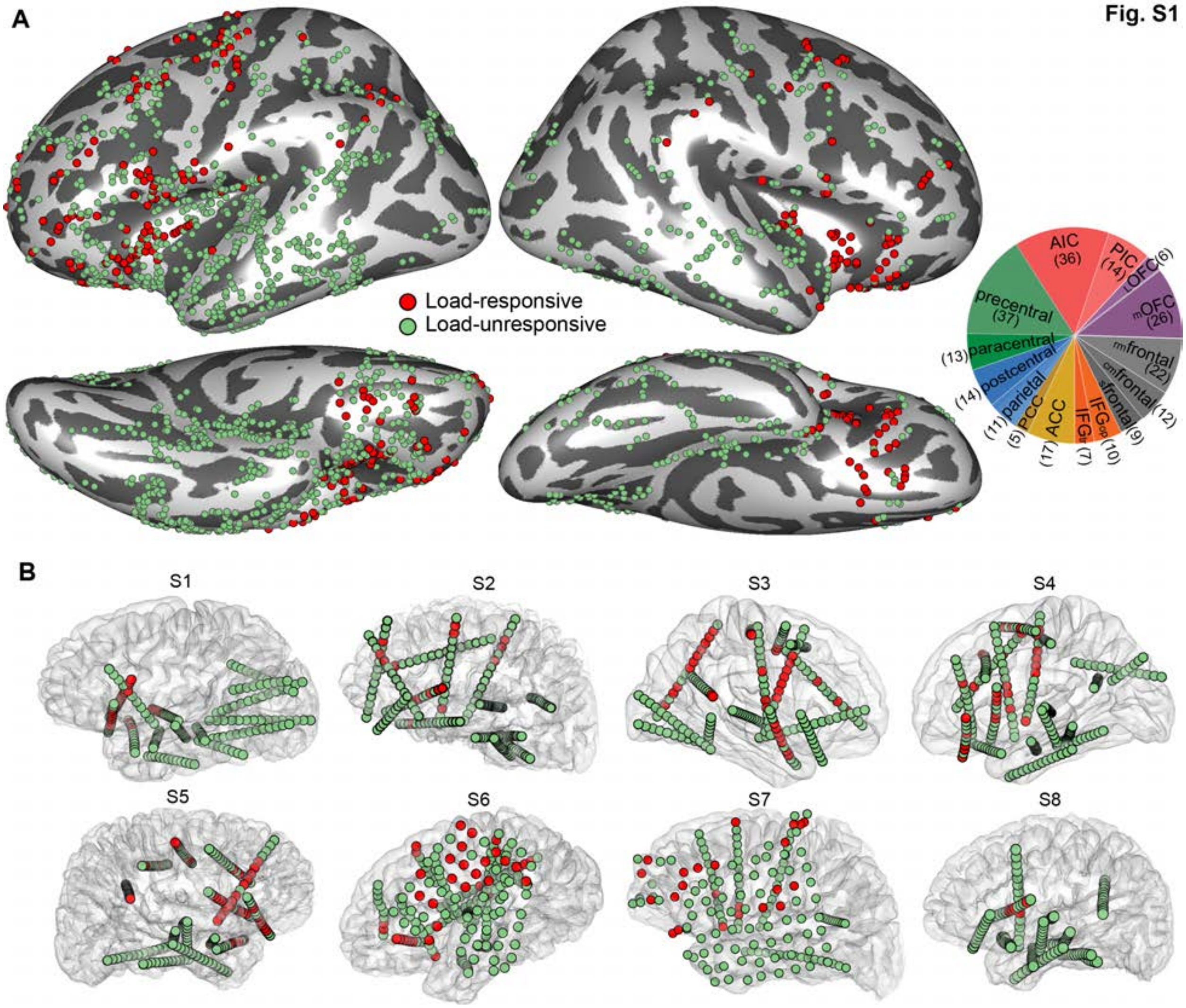


**Figure S1. Electrode coverage and load effects.**
**(A)** Load-responsive (red) and unresponsive (green) electrodes from all 8 participants overlaid on the FreeSurfer average inflated surfaces. Of 1328 electrodes, 239 (18%) showed significant load responses (greater HFA in loaded vs. preloaded inhalations, 0–3 s post-inhalation onset). Pie chart shows the regional distribution of these 239 load-responsive electrodes; numbers in brackets indicate the total number of electrodes sampled in each brain region.
**(B)** Same as A, but shown separately for each participant. Translucent pial surface plots display both epidural and deeper electrodes on the individual brain. Most representative hemisphere is shown for each participant. Participants 3 and 4 had some electrodes in the contralateral hemisphere (not shown). Participants 6 and 7 were implanted with both depths and grids. Electrode contacts are enlarged for visibility.

Fig. S2

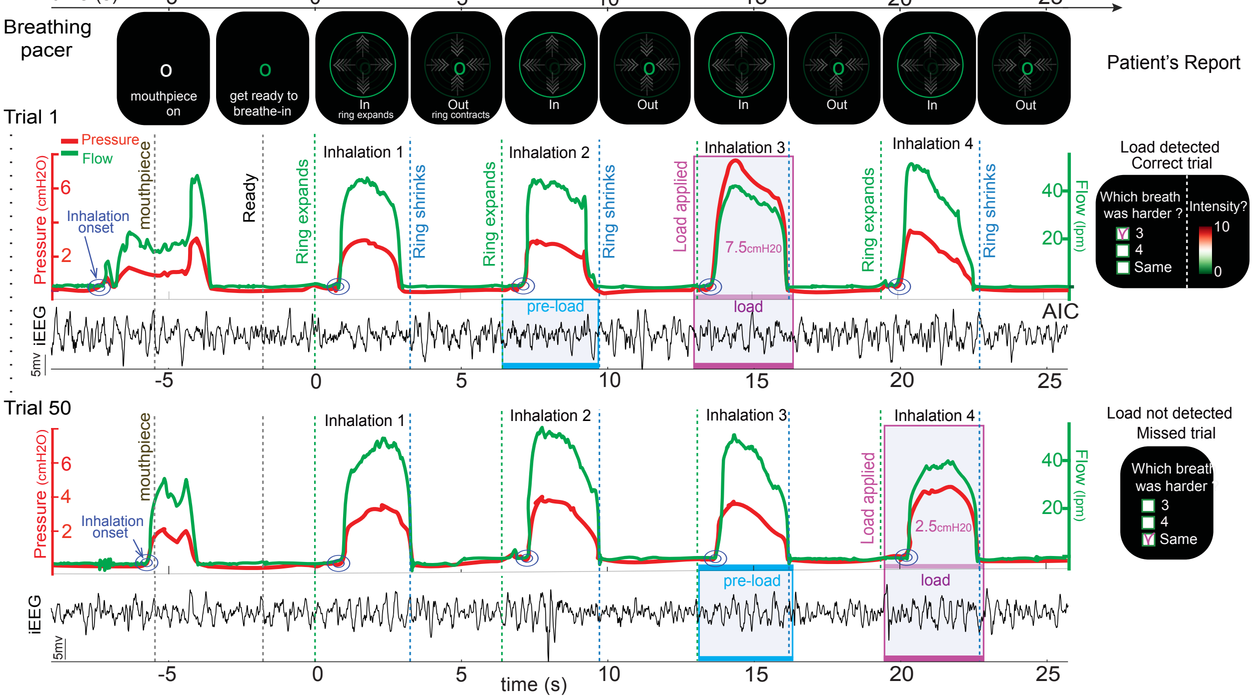


**Figure S2. Example RRST trials with respiratory and neural recordings.**
Two example trials from the Respiratory Resistance Sensitivity Task (RRST), where patients took four paced breaths (3 s inhale, 3 s exhale) guided by an expanding/contracting ring (green/blue dashed lines) and reported if a load was applied and its intensity. Loads were unpredictably delivered on one inhalation per trial. Traces show inspiratory airflow (iAF), mouth pressure (iMP), and HFA from an anterior insular cortex (AIC) electrode. Top: detected 7.5 $cmH_2O/L/s$ load. Bottom: undetected 2.5 $cmH_2O/L/s$ load. Blue dots mark inhalation onsets.

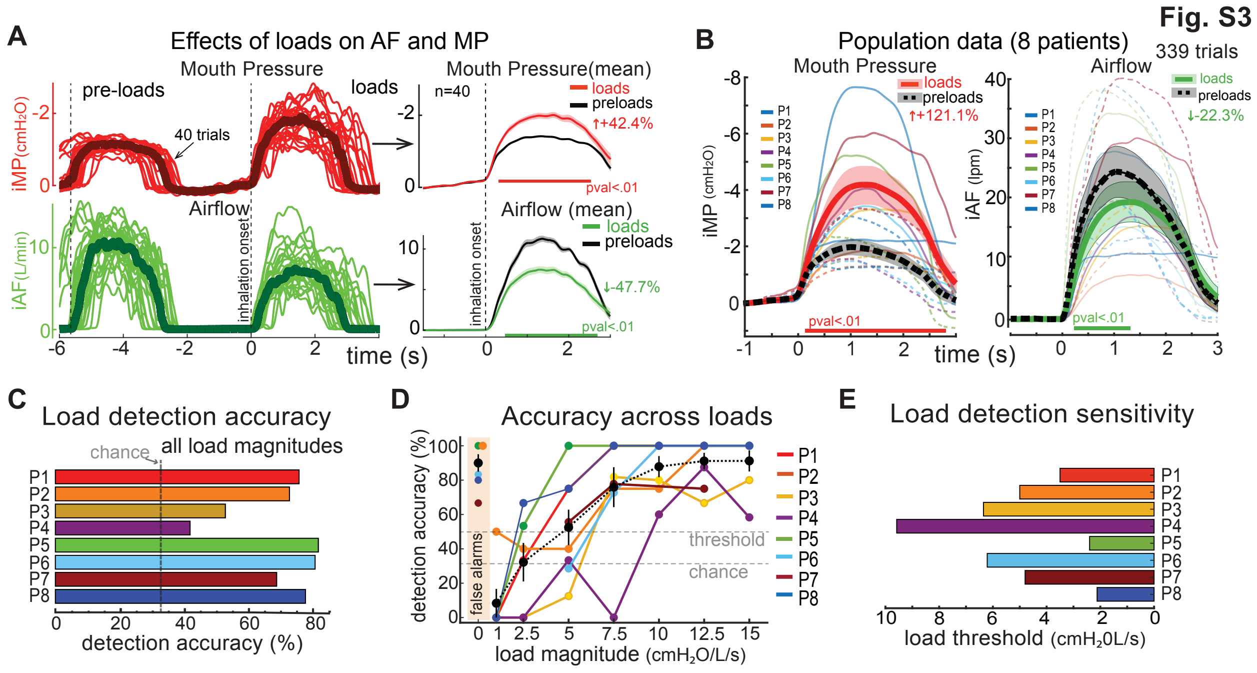


**Figure S3. Load-dependent changes in inspiratory effort and detection accuracy.**
**(A)** Inspiratory mouth pressure (iMP, red) and airflow (iAF, green) from an example participant across 40 trials (7 load magnitudes). iMP rose and iAF fell during loaded vs. preloaded inhalations (0–3 s post-onset). Thin lines show single trials; thick lines show means. Right: overlaid mean traces ± SEM.
**(B)** Group iMP (left) and iAF (right) across all participants (n = 8; 339 trials). Thin colored lines indicate participant means; thick dashed lines show grand averages.
**(C)** Overall detection accuracy. Bars indicate percent correctly detected trials across all loads; dashed line marks chance (33%).
**(D)** Detection accuracy by load magnitude for each participant.
**(E)** Detection thresholds, defined as the lowest load detected >50% of the time.

**Fig. S4**

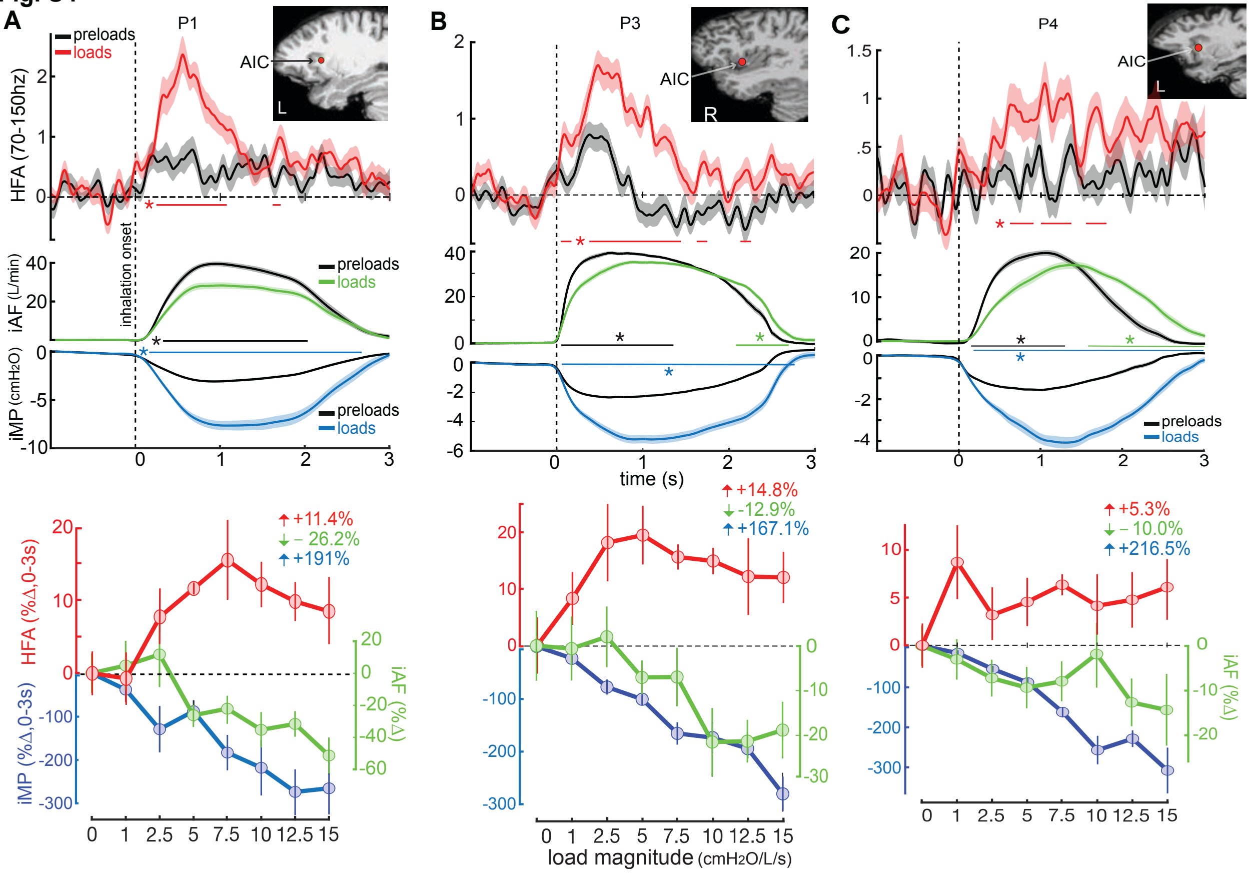


**Figure S4. Load-responsive electrodes in the anterior insular cortex (AIC).**

Three representative AIC electrodes from different participants showing load-evoked neural and respiratory responses.

(A) Left AIC in participant S1 (48 trials).

(B) Right AIC in participant S3 (42 trials).

(C) Left AIC in participant S4 (38 trials).

Each panel shows (left) time courses of HFA (70–150 Hz; top), inspiratory airflow (iAF; green), and mouth pressure (iMP; blue) averaged across loaded (red) vs. preloaded (black) inhalations, and (right) mean responses as a function of load magnitude (0–3 s post-inhalation). HFA rose with load alongside increased inspiratory effort (iMP, iAF). Asterisks indicate significant differences from preloads ($p < 0.05$, ranksum). Shaded areas denote SEM. Vertical dashed lines mark inhalation onset.

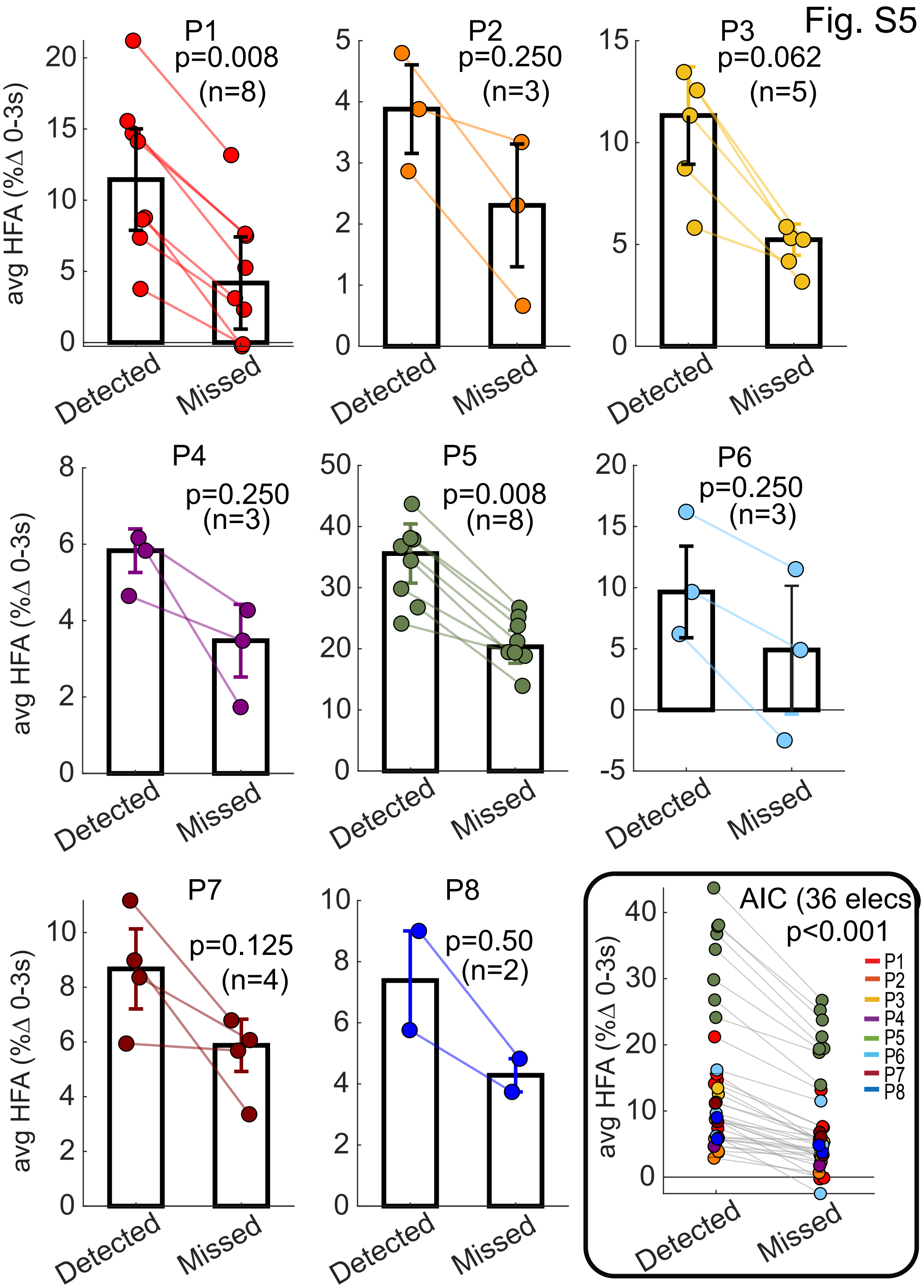


**Figure S5. Individual AIC responses to detected versus missed loads.**
Mean HFA gains (0–3 s post-inhalation) in the anterior insular cortex (AIC) for detected vs. missed trials, plotted separately for each participant (n = 8). Trials were matched by load magnitude. Bars show mean ± SEM. (Inset) Group-averaged HFA gains across all AIC electrodes (n = 36).

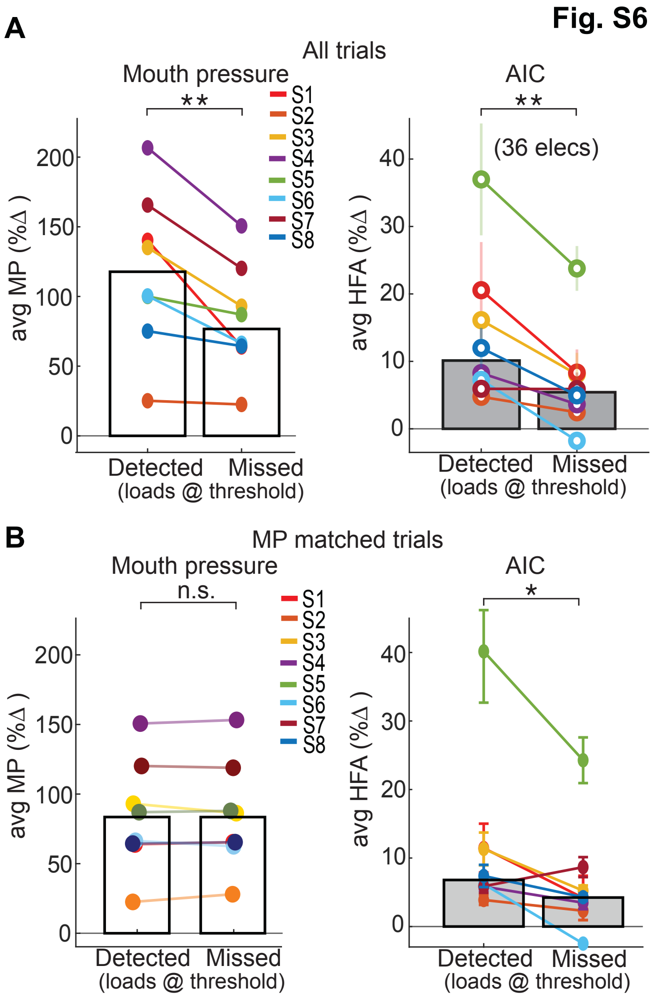


**Figure S6. Mouth pressure differences in detected vs. missed trials.**
**(A)** All trials: Inspiratory mouth pressure (iMP, left) was significantly higher for detected versus missed trials of the same magnitude ($p = 0.008$; Wilcoxon signed-rank), while AIC HFA gains (right, 36 electrodes) were also significantly greater in detected trials.
**(B)** After matching trials to control for iMP (no significant pressure difference), HFA gains in the AIC remained higher for detected loads of the same magnitude ($p = 0.039$), indicating that increased respiratory effort did not fully explain neural differences. Each line shows data from one participant (colors), bars indicate group mean ± SEM.

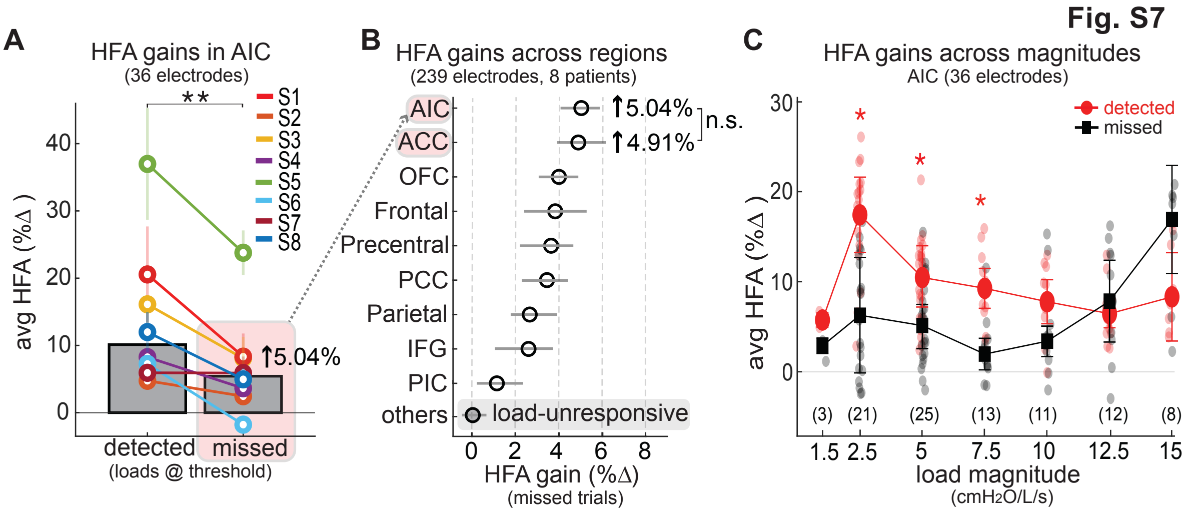


**Figure S7. Neural responses during missed load trials.**
Even without detection, modest AIC HFA gains indicate subthreshold processing.
**(A)** AIC electrodes showed small but significant HFA increases (~5%) during missed trials.
**(B)** These effects were most prominent in the AIC and ACC.
**(C)** HFA gains in detected trials peaked at lower loads (where detection is critical), while missed-trial gains increased more for higher loads. Numbers denote electrode counts per magnitude.
**Abbreviations:** HFA = high-frequency activity; OFC = orbitofrontal cortex; PCP = postcentral/parietal cortex; SMA = supplementary motor area; PMC = premotor cortex; ACC = anterior cingulate cortex; AIC = anterior insular cortex.

Fig. S8

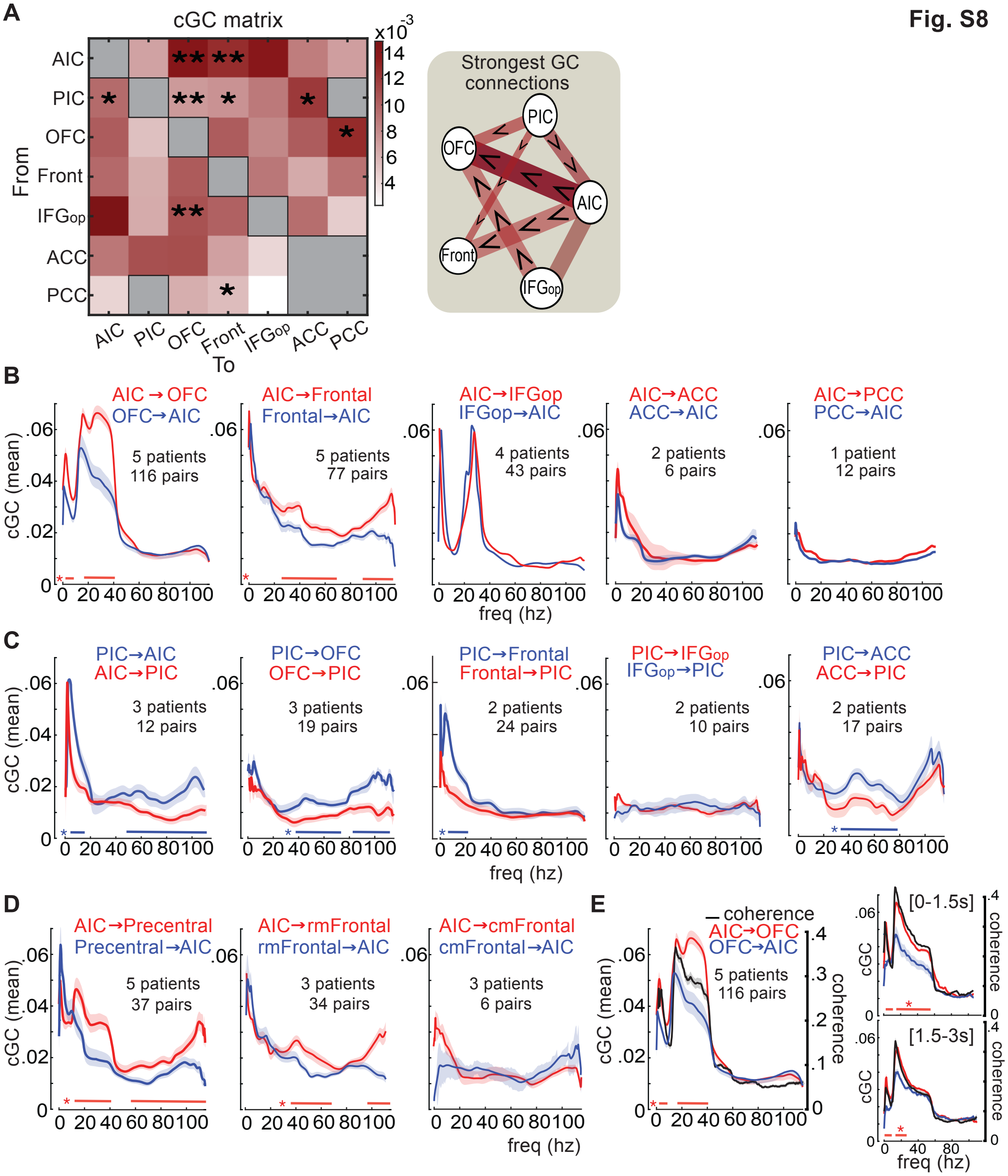


**Figure S8. Connectivity profiles supporting load detection.**
**(A)** Conditional Granger causality (cGC) matrix across load-responsive brain regions during loaded trials (0–3 s post-inhalation). Darker red indicates a larger absolute GC estimate; asterisks mark where the GC in one direction is significantly higher than the reverse, indicating directional asymmetry. Right: schematic summarizes dominant connections (arrow direction, line width/color denote strength).
**(B)** Spectral-resolved cGC from AIC to target regions, showing a distinct peak at 15–40 Hz for AIC→OFC and a broader elevation spanning 25–115 Hz for AIC→frontal cortex. Asterisks along the x-axis indicate frequency bins where GC directionality (AIC→target vs. reverse) was

significant. GC was computed for each electrode pair (e.g., AIC#1 and OFC#1 in P1, AIC#1 and OFC#2 in P1, and so on across all participants) and then pooled to generate these curves.
**(C)** Spectral-resolved cGC from PIC, showing broader but weaker influence than AIC.
(D) AIC→frontal cGC separated by subregion, strongest for precentral cortex.
**(E)** AIC→OFC coherence peaked at 18 Hz, matching GC frequencies, and was reduced (but not absent) in later inhalation phases (1.5–3 s vs. 0–1.5 s).

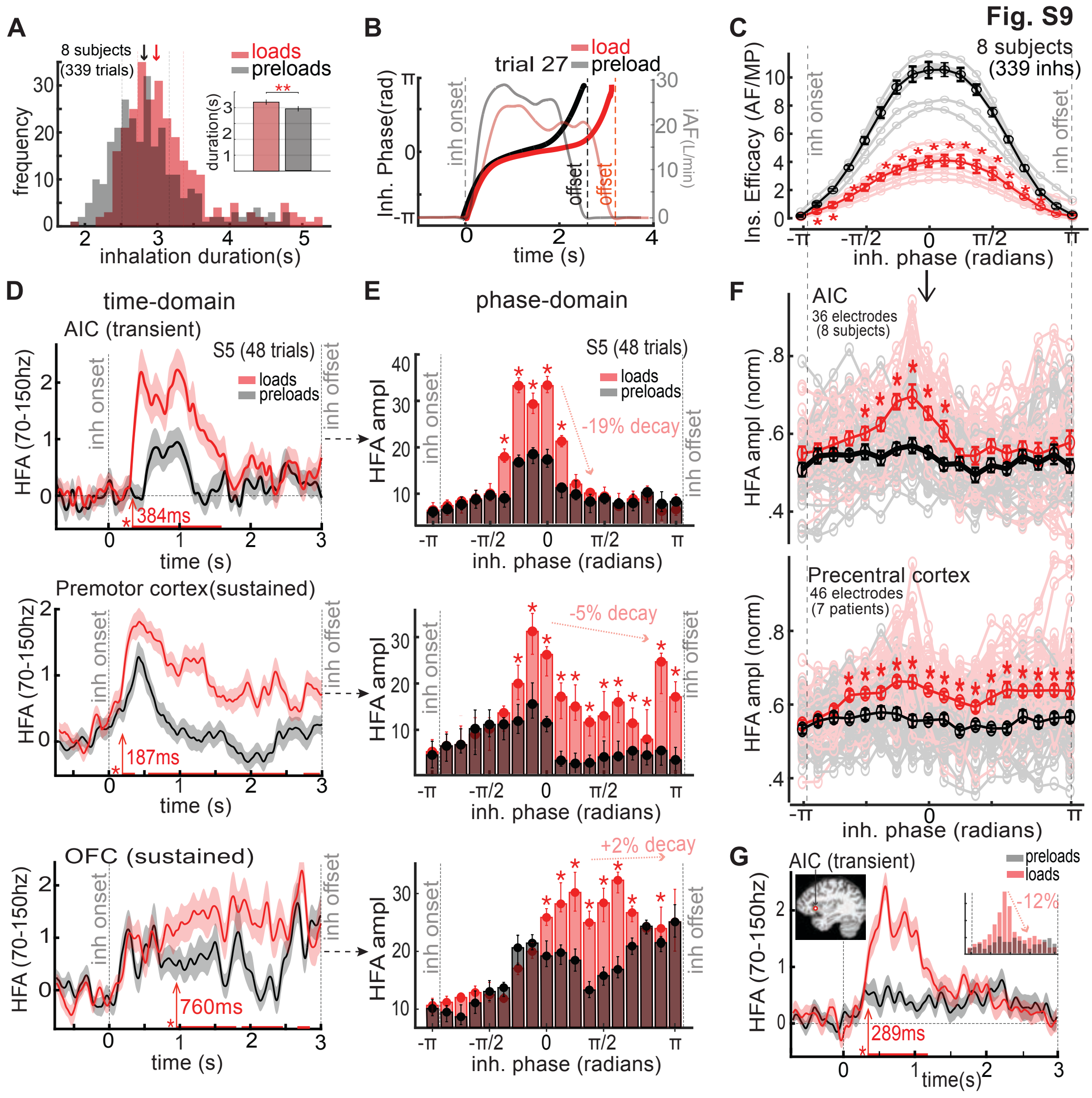


**Figure S9. Transient vs. sustained load responses and phase-aligned HFA dynamics across cortical regions.**

**(A)** Inhalation durations for all preloaded and loaded trials (n = 339; 8 participants). Preloaded inhalations were slightly shorter ($p < 0.01$); inset shows group means ± 95% CI.

**(B)** Example trial showing conversion of airflow to inhalation phase. Each inhalation was normalized from 0–360° (–π to π radians) to account for duration variability.

**(C)** Phase-aligned inspiratory effort index (iAF/iMP) across all participants and trials. Each line shows the averaged IE for one participant. Preloaded trials showed reduced effort, peaking near mid-inhalation. Asterisks indicate significant bin differences.

**(D)** Time-domain HFA (70–150 Hz) responses from AIC, premotor, and OFC electrodes in one participant (S5; 48 trials, 7 load magnitudes). AIC responses were transient, whereas frontal regions showed sustained activity. Red arrows mark load-related response latencies.

**(E)** Phase-domain HFA from the same electrodes. AIC showed brief early increases, while premotor and OFC responses extended across the inhalation. Each bin shows the median and interquartile range across trials. Asterisks indicate significant bins (signed-rank, $p < 0.05$).

**(F)** Population phase-binned HFA for all AIC ($n = 36$) and precentral ($n = 46$) electrodes. Asterisks mark significant bins ($p < 0.05$).
**(G)** Time- and phase-domain HFA for the AIC electrode shown in Fig. 2B.

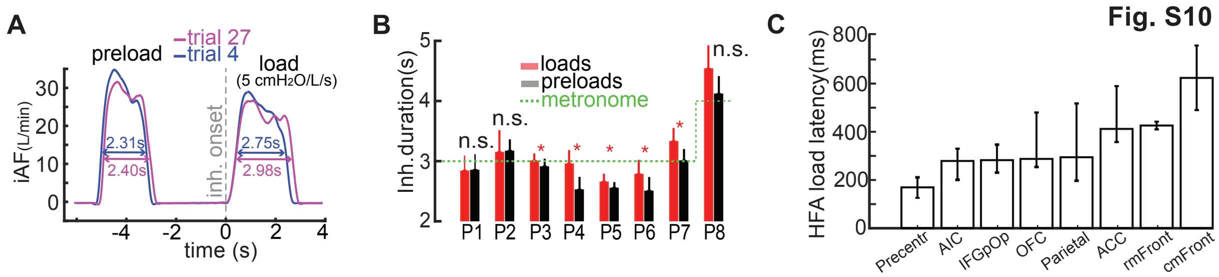


**Figure S10. Breath-to-breath variability and latency of neural responses.**
**(A)** Example trials showing breath-to-breath variability in inhalation duration across preloaded and loaded conditions.
**(B)** Inhalation durations by participant and condition. Bars indicate mean ± SE across trials. Dashed green line marks the pacer target (3 s inhalations for all participants except Participant 8, who used 4 s to better match her natural breathing pace).
**(C)** Latency of load-related HFA increases across cortical regions. Bars show median onset latency for significant HFA elevations (load vs. preload). Motor areas (precentral, including motor, premotor, and SMA) responded earlier than AIC and OFC, suggesting rapid motor engagement during inspiratory load processing. See Fig. S9D for single-electrode examples.

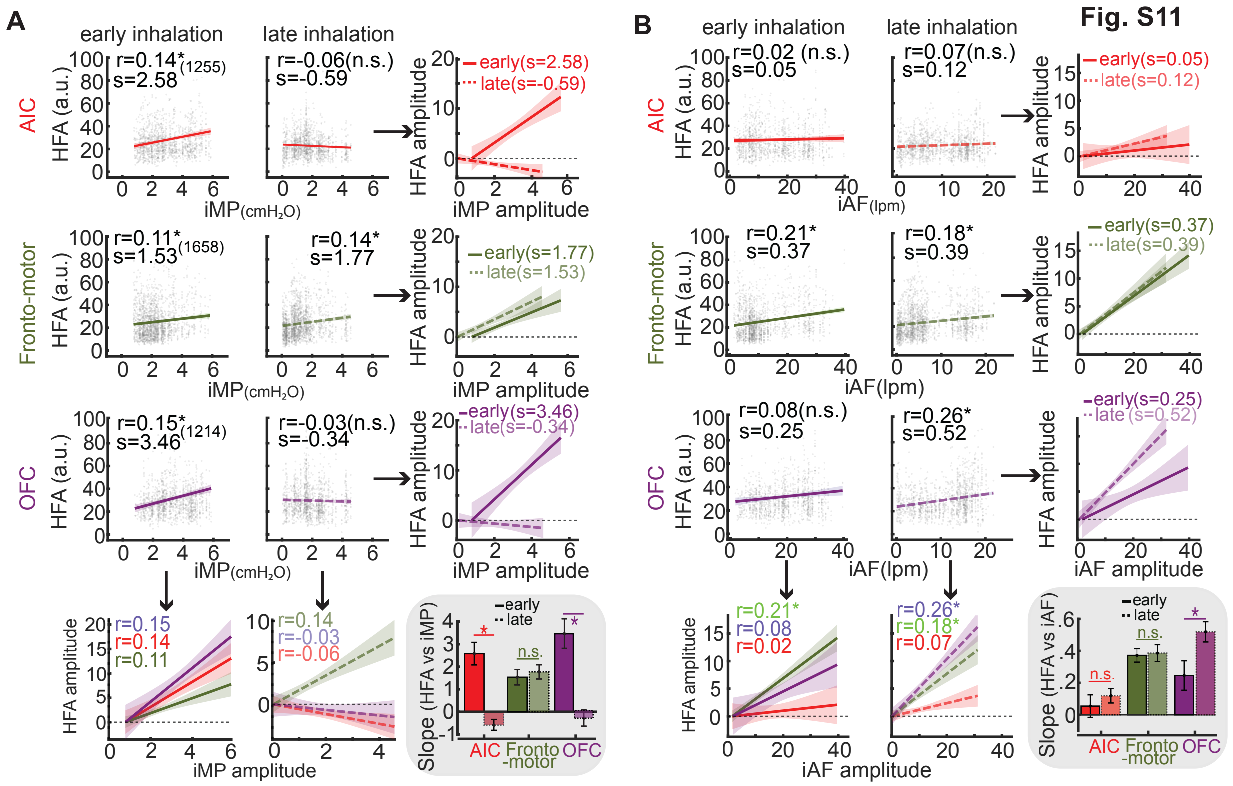


**Fig. S11. Distinct cortical encoding of inspiratory pressure and airflow across inhalation phases.**

**(A)** Trial-level correlations between high-frequency activity (HFA) and inspiratory mouth pressure (iMP) amplitude were computed during early and late inhalation in the anterior insula (AIC), fronto-motor cortex, and orbitofrontal cortex (OFC). The AIC selectively encoded pressure in the early phase. Fronto-motor regions showed significant correlations in both phases. OFC exhibited prominent early-phase sensitivity to pressure. Each subplot shows Pearson correlation (r), slope (s), and individual trial data. In plots with overlaid regression lines (arrows), HFA was aligned to zero at the minimum iMP to facilitate slope comparison. Summary plots (grey box) compare slope differences across regions and phases. Asterisks indicate significant Pearson correlations or significant slope differences based on signed-rank tests after FDR correction.

**(B)** Same analysis as in (A), using inspiratory airflow (iAF). Fronto-motor regions again showed robust, phase-invariant modulation. The AIC exhibited weak airflow encoding. OFC responses were minimal in the early phase but increased in the late phase, with higher correlation and slope values. Summary plots (grey box) illustrate signal- and phase-specific encoding across regions.

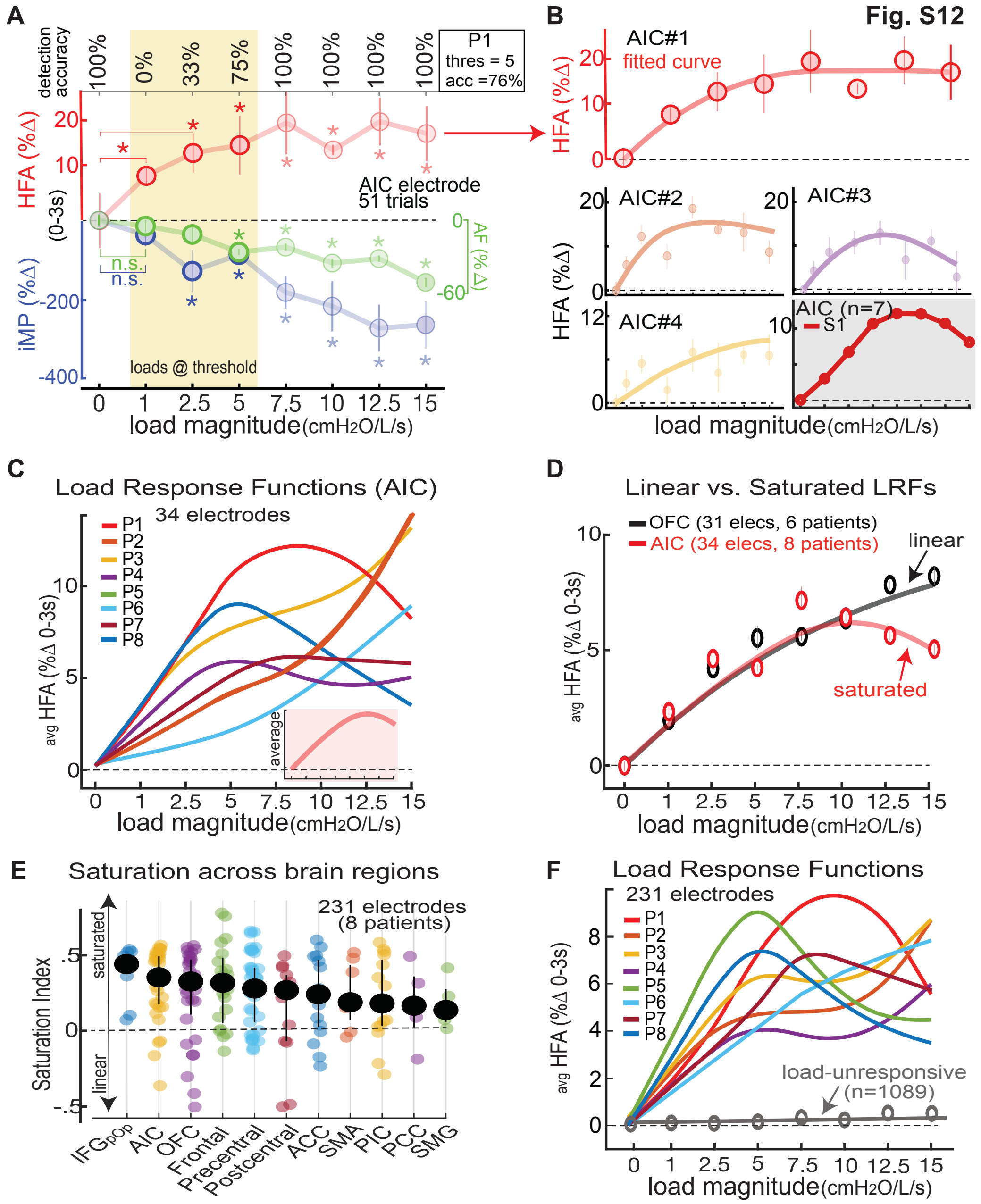


**Fig. S12. Load-magnitude tuning and saturation across cortical regions.**
**(A)** Same AIC electrode as Fig. 2B, showing HFA (red), inspiratory pressure (iMP, blue), and airflow (iAF, green) across increasing load magnitudes. HFA rose up to ~10 cmH₂O/L/s, then plateaued or declined. Detection accuracies are marked above.
**(B)** Top: Curve fit to mean HFA from this electrode. Bottom: Load-magnitude response functions (LRFs) from three AIC electrodes in the same participant, all showing saturation. Right: Averaged LRF across all seven AIC electrodes from this participant (one electrode failed goodness-of-fit and was excluded).
**(C)** LRFs from AIC electrodes across eight participants (34/36 electrodes passed the goodness-of-fit test).

**(D)** Participant-averaged LRFs in AIC and OFC. AIC responses saturated early, whereas OFC responses increased more linearly.
**(E)** Saturation index by region. Each dot represents one electrode; black circles show regional medians ± IQR. AIC and IFGop showed the strongest saturation.
**(F)** Load-magnitude response profiles (LMRs) from all significantly load-responsive electrodes ($n = 231$; eight electrodes excluded for poor fit). Most showed plateauing or declining HFA at higher loads. Dashed gray line shows the average response across load-unresponsive electrodes ($n = 1089$).

| ptID | Sex | Brain Anomaly | SOZ | Elecs | SOZ - spread |
|---|---|---|---|---|---|
| P1 | M | TO heterotopia | L sup P/O | 234 (L/R) | Spread to L hippocampus |
| P2 | F | none | L mid-sup T | 280 (R) | focal |
| P3 | F | Multifocal encephalopathy | R T operculum | 153 | R O/P |
| P4 | M | none | L amygdala | 249 | hippocampus |
| P5 | M | none | R mesial T lobe | 215 | T pole, OFC |
| P6 | F | cortical dysplasia | multifocal | 167 | Perisylvian |
| P7 | M | none | L lateral T | 223 | L F |
| P8 | F | none | L mid/inf T | 237 | L Mesial T |
| P9* | F | R temporal cavernoma | R Insula | 218 | R T operculum |
| P10* | M | L post T cavernoma | L mes T | 161 | OFC |
| P11* | F | none | L hippocampal | 216 | L T tip |

**Table S1: Patient seizure information**
T = temporal; O = occipital; P = Parietal. F = Frontal. Electrodes = total electrodes implanted across both hemispheres; L = left hemisphere; R = right hemisphere; s = superior (e.g., sP = superior parietal); SOZ = seizure onset zone; Sz = seizures. Asterisk (*) indicates patients excluded from main analyses due to task non-compliance or excessive epileptogenic discharges within targeted (interoceptive) brain regions.

| | FEV1 (L) | FVC (L) | FEV1/FVC (%) | Raw (cmH2O/L/s) | %Pred FEV1 | %Pred FVC |
|---|---|---|---|---|---|---|
| P1 | 3.7 | 4.1 | 83.2 | 3.6 | 96.4 | 100.2 |
| P2 | 3.6 | 4.5 | 84.8 | 4.2 | 92.2 | 97.7 |
| P3 | 3.8 | 4.1 | 83.4 | 3.1 | 99.7 | 96.4 |
| P4 | 4 | 4.1 | 82.8 | 4.4 | 91.6 | 95.6 |
| P5 | 3.6 | 4.4 | 86.1 | 3.5 | 97.3 | 90.4 |
| P6 | 3.6 | 3.6 | 84.2 | 3.8 | 88.6 | 93.8 |
| P7 | 4 | 3.6 | 84.5 | 3.5 | 91.1 | 94.9 |
| P8 | 3.8 | 4.1 | 82.8 | 5.7 | 97.2 | 101.6 |

**Table S2. Pulmonary function tests.**
Pulmonary function tests conducted prior to the RRST task. $FEV_1$, forced expiratory volume in 1 s; FVC, forced vital capacity; Raw, airway resistance (measured via impulse oscillometry). Values reflect the highest of three FVC trials per participant. All participants had $FEV_1$ > 70% of predicted, indicating normal respiratory function (42).
